# Impact of age-structure and vaccine prioritization on COVID-19 in West Africa

**DOI:** 10.1101/2022.07.03.22277195

**Authors:** Hemaho B. Taboe, Michael Asare-Baah, Afsana Yesmin, Calistus N. Ngonghala

**Author notes:** Corresponding author: Calistus N. Ngonghala.

## Abstract

The ongoing COVID-19 pandemic has been a major global health challenge since its emergence in 2019. Contrary to early predictions that sub-Saharan Africa (SSA) would bear a disproportionate share of the burden of COVID-19 due to the region’s vulnerability to other infectious diseases, weak healthcare systems, and socioeconomic conditions, the pandemic’s effects in SSA have been very mild in comparison to other regions. Interestingly, the number of cases, hospitalizations, and disease-induced deaths in SSA remain low, despite the loose implementation of non-pharmaceutical interventions (NPIs) and the low availability and administration of vaccines. Possible explanations for this low burden include epidemiological disparities, under-reporting (due to limited testing), climatic factors, population structure, and government policy initiatives. In this study, we formulate a model framework consisting of a basic model (in which only susceptible individuals are vaccinated), a vaccine-structured model, and a hybrid vaccine-age-structured model to reflect the dynamics of COVID-19 in West Africa (WA). The framework is trained with a portion of the confirmed daily COVID-19 case data for 16 West African countries, validated with the remaining portion of the data, and used to (i) assess the effect of age structure on the incidence of COVID-19 in WA, (ii) evaluate the impact of vaccination and vaccine prioritization based on age brackets on the burden of COVID-19 in the sub-region, and (iii) explore plausible reasons for the low burden of COVID-19 in WA compared to other parts of the world. Calibration of the model parameters and global sensitivity analysis show that asymptomatic youths are the primary drivers of the pandemic in WA. Also, the basic and control reproduction numbers of the hybrid vaccine-age-structured model are smaller than those of the other two models indicating that the disease burden is overestimated in the models which do not account for age-structure. This result is also confirmed through the vaccine-derived herd immunity thresholds. In particular, a comprehensive analysis of the basic (vaccine-structured) model reveals that if 84% (73%) of the West African populace is fully immunized with the vaccines authorized for use in WA, vaccine-derived herd immunity can be achieved. This herd immunity threshold is lower (68%) for the hybrid model. Also, all three thresholds are lower (60% for the basic model, 51% for the vaccine-structured model, and 48% for the hybrid model) if vaccines of higher efficacies (e.g., the Pfizer or Moderna vaccine) are prioritized, and higher if vaccines of lower efficacy are prioritized. Simulations of the models show that controlling the COVID-19 pandemic in WA (by reducing transmission) requires a proactive approach, including prioritizing vaccination of more youths or vaccination of more youths and elderly simultaneously. Moreover, complementing vaccination with a higher level of mask compliance will improve the prospects of containing the pandemic. Additionally, simulations of the model predict another COVID-19 wave (with a smaller peak size compared to the Omicron wave) by mid-July 2022. Furthermore, the emergence of a more transmissible variant or easing the existing measures that are effective in reducing transmission will result in more devastating COVID-19 waves in the future. To conclude, accounting for age-structure is important in understanding why the burden of COVID-19 has been low in WA and sustaining the current vaccination level, complemented with the WHO recommended NPIs is critical in curbing the spread of the disease in WA.

## 1. Introduction

Challenges associated with the 2019 coronavirus (COVID-19) pandemic have been enormous. The pandemic has impacted every country and region of the world with over 543 million reported cases and 6.3 million deaths as of late June 2022 [1]. Although its impact on the African continent has not been as devastating as in other parts of the globe or as predicted earlier, over 11.7 million confirmed cases and 253, 935 deaths had been recorded in Africa and 847, 400 cases and 11, 400 deaths in the West African sub-region as of June 22, 2022 [2]. In the West African sub-region, the pandemic has been aggravated by an already weakened economy, healthcare system, and immunity among a large proportion of the population due to the high prevalence of malnutrition, and other acute and chronic infectious diseases [3]. Specifically, poor socio-economic conditions coupled with the overwhelming population density that depends largely on face-to-face interactions for their basic livelihoods challenges the practicality of non-pharmaceutical interventions (NPIs) such as social distancing, self-isolation of confirmed cases, quarantining of suspected cases, and nationwide lockdown and travel ban measures [4]. The secondary ramifications of imposing these measures with strict compliance can impact the survival and livelihoods of these populations negatively, with a potential increase in morbidity and mortality arising from hunger, financial difficulties and non-utilization of essential services such as primary health care [4–6].

Notwithstanding these challenges, sub-Saharan Africa (SSA) appears to be minimally impacted by the pandemic, with fewer confirmed and reported COVID-19 cases and mortalities than elsewhere. The demographic distribution (a key epidemiological indicator of disease transmission), low case detection rate due to insufficient testing capacity, epidemiological disparities, and timely implementation of NPIs best explain the variations in the burden of COVID-19 observed across different settings [7–10]. The young population age-structure in Africa with a median age of 19 years is significant in limiting morbidity and mortality associated with the disease [7, 8]. Evidence from various studies have associated a greater risk of disease severity and mortality with older and immunocompromised individuals [11–13]. Increased resistance to infections can be acquired through enhanced immunity from vaccination or cross-protection from previous infections [14]. The latter is more conceivable among younger individuals who are more likely to be exposed to other coronaviruses and respiratory viruses [11, 12, 15]. Moreover, the younger population is more likely to experience milder (asymptomatic or paucisymptomatic) disease conditions, which are rarely noticed or reported [11], though, they remain transmissible. The high prevalence of subclinical infections among young people justifies the low number of reported clinical cases, hospitalizations, and mortality in populations with significant proportions of younger individuals, as observed in many low-income countries [13]. Younger people tend to have a broader network of social contacts than the elderly and contribute substantially to the transmission of the pathogen. Due to their milder and subclinical disease nature, most of them are usually unaware of their disease status [16]. On the contrary, elderly with fewer interaction networks and more cautious nature bear the brunt of clinical disease, hospitalization, and death.

Considering the socio-economic structure and population demographics of most countries in West Africa (WA), effective pharmaceutical interventions (vaccines and anti-viral medications) stand out as the strategy of choice to reduce transmission and/or curtail the burden of the pandemic [17]. Conventionally, the tenet of vaccination can be achieved by directly vaccinating individuals at high risk of severe disease, hospitalization, and death and/or protecting these vulnerable groups by vaccinating those primarily responsible for transmitting the pathogen [18]. The WHO authorized COVID-19 vaccines presently in use in Africa include AstraZeneca, Sinopharm, Sputnik V, BioNTech, Sinovac, Covaxin, Sputnik Light, Pfizer, and Moderna [19]. These vaccines effectively meet the tenet of vaccination explained above, although with varying efficacy ranging between 51% - 95% [20]. As of June 28, 2022, 72% of vaccines doses supplied in Africa have been administered, with 18% of the population fully vaccinated since the commencement of the vaccination program in December 2020 [19]. The low vaccination intake can be attributed to vaccine hesitancy arising from misinformation and risk perception of available vaccines [21, 22]. Despite efforts to address this challenge, there remain a gap in the quantity of vaccine supplied. The limited availability of COVID-19 vaccines in most low-income countries, mainly obtained from the COVAX assistance program or donor supports [23], calls for the prioritization of vaccine administration to achieve its maximum impact. Understanding the influence of the population age structure on the transmission and incidence of COVID-19 in WA will ensure efficient utilization of resources for maximum impact.

Mathematical models have provided comprehensive insight into COVID-19 dynamics and informed various interventions [9, 24–30]. These models have been instrumental in developing effective policies that have steered public health directions and effected change in many settings. A study that deployed an age-structured model fitted to COVID-19 epidemic data from China, Italy, Japan, Singapore, Canada, and South Korea [11] showed a significant increase of 57–82% in clinical infections among individuals 70 years and over. In a similar study in [23], the authors demonstrated that vaccinating not just those above 60 years but also those from 5 – 60 years reduced disease incidence substantially. Findings from [31] using an age-structured epidemiological model proved that regardless of vaccine efficacy, control measures and immunity dynamics, prioritizing vaccine administration among the elderly (> 60 years) had the highest relative reduction in deaths. Although prioritizing vaccination of the adult population will reduce severe disease and mortality, focusing on younger people can reduce transmission and decrease the incidence of the disease [32, 33]. These studies justify prioritizing the more aging population in high-income countries during the initial phase of vaccine allocation.

Here, we formulate and use a mathematical framework that accounts for vaccination and age-structure to assess the contributions of youths and elderly to COVID-19 prevalence in WA. The framework is used to evaluate the impact of vaccination and vaccine prioritization based on age brackets on the burden of COVID-19 in the sub-region and to explore possible reasons why the burden of the COVID-19 pandemic has been less severe in WA compared to other regions.

## 2. Methods

Three compartmental models depicting the transmission dynamics of COVID-19 in the West African sub-region are developed. The first model (referred to as Model 1 or the basic model) accounts for vaccine-derived immunity and the waning effects of vaccinated and recovered individuals. The second model (referred to as Model 2)–an extension of the first model, caters for the heterogeneity in vaccination status; it is structured based on whether individuals in each of the classes are vaccinated or unvaccinated. The third model (referred to as Model 3) is an extension of the second model that accounts for age structure, with individuals subdivided into two age groups determined by disease risk. In all three models, vaccinated individuals are those who have been fully vaccinated (i.e., individuals who have received all required vaccine doses, including booster doses).

### 2.1. Model 1: The basic model

The model, that is based on an extended Kermack-Mckenderick type framework [34] splits the entire human population into nine distinct groups based on disease and vaccination status. The classes include unvaccinated susceptible (*S*_*u*_), vaccinated susceptible (*S*_*v*_), latent (*E*), presymptomatic infectious (*I*_*p*_), symptomatic infectious (*I*_*i*_), asymptomatic infectious (*I*_*a*_), confirmed cases (*I*_*c*_), hospitalized (*I*_*h*_), and recovered (*R*). Hence, the total population denoted by *N* is

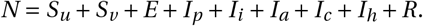

The model (2.1), assumes no vertical transmission of COVID-19, hence, births from all the classes enter into the unvaccinated susceptible class (*S*_*u*_) at rate Λ. Natural deaths in each class occur at per capita rate *µ*. Vaccination of susceptible individuals is at per capita rate *ξ*, while vaccine-derived immunity of vaccinated susceptible individuals wanes at per capita rate *ω*_*sv*_, and natural immunity of recovered individuals wanes at per capita rate *ω*_*r*_. Unvaccinated (vaccinated) susceptible individuals become infected and progress to the latent class (*E*) at per capita rate *λ* = (*β*_*p*_ *I*_*p*_ + *β*_*a*_ *I*_*a*_ + *β*_*i*_ *I*_*i*_ + *β*_*c*_ *I*_*c*_ + *β*_*h*_ *I*_*h*_)/*N* ((1 − *ε*)*λ*), where *β*_*j*_, *j* ∈ {*p, a, i, c, h*} is the effective contact rate for members of the *I* _*j*_ th class and 0 ≤ *ε* ≤ 1 is the efficacy of vaccines in preventing vaccinated individuals from contracting the virus. Latent individuals progress to the presymptomatic infectious class at per capita rate *σ*_*e*_, while at the end of the incubation period, presymptomatic infectious individuals join the symptomatic (asymptomatic) infectious class at per capita rate *rσ*_*p*_ ((1 − *r*)*σ*_*p*_), where 0 ≤ *r* ≤ 1 is the fraction of presymptomatic infectious individuals who develop clinical disease symptoms. The confirmed class is populated by detected individuals (i.e., individuals who test positive) from the latent, presymptomatic infectious, and asymptomatic infectious classes at per capita rate, *τ*_*a*_, and detected individuals from the symptomatic infectious class at per capita rate, *τ*_*i*_. It should be mentioned that in WA, apart from travel and administrative reasons, most of the people who are tested are the symptomatic infectious. Therefore, *τ*_*i*_ > *τ*_*a*_. Confirmed (symptomatic infectious) individuals are hospitalized at per capita rate *ϕ*_*c*_ (*ϕ*_*i*_), while confirmed, symptomatic infectious, asymptomatic infectious, and hospitalized individuals recover from infection at per capita rate, *γ*_*c*_, *γ*_*i*_, *γ*_*a*_, and *γ*_*h*_, respectively. COVID-19 related deaths occur in the confirmed, symptomatic infectious, and hospitalized classes at per capita rate, *δ*_*c*_, *δ*_*i*_, and *δ*_*h*_, respectively. Descriptions of the variables and parameters of the basic model (2.1) are summarized in Tables S1-S2 of the supplementary information (SI), while the model is depicted schematically in Fig. 1.

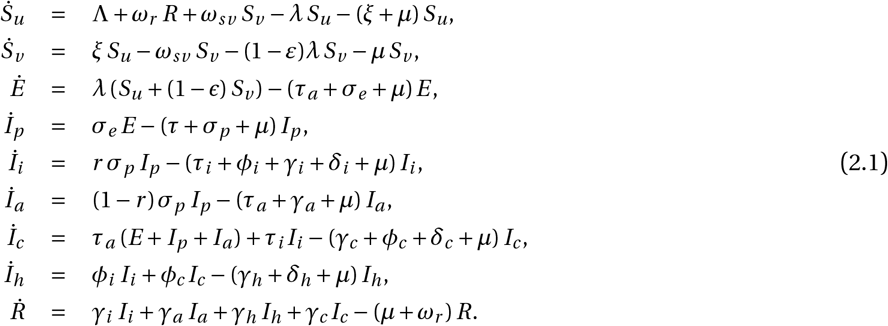

**Fig. 1:**
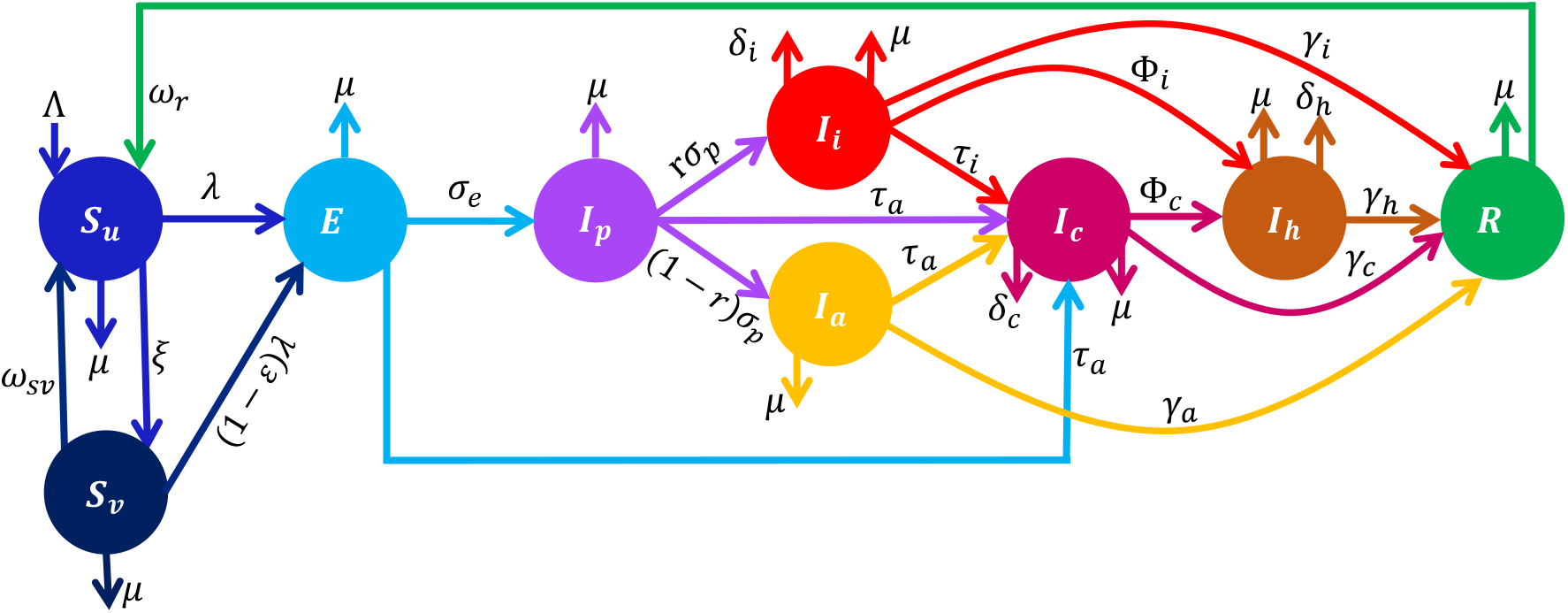
Schematic diagram of the basic model depicting the flow of individuals between classes based on the disease and vaccination status. The classes are unvaccinated susceptible (*S*_*u*_), vaccinated susceptible (*S*_*v*_), latent (*E*), presymptomatic infectious (*I*_*p*_), symptomatic infectious (*I*_*i*_), asymptomatic infectious (*I*_*a*_), confirmed infectious (*I*_*c*_), hospitalized (*I*_*h*_), and recovered (*R*). Descriptions of the model variables and parameters are provided in Table S1-S2 of the SI.

### 2.2. Model 2: The vaccine-structured model

The vaccine-structured model is derived by splitting the population into non-vaccinated and vaccinated groups (with subscripts *u* and *v*, respectively). Hence, the total population (*N*) is *N* = *N*_*u*_ + *N*_*v*_, where *N*_*u*_ is the total non-vaccinated population and *N*_*v*_ is the total vaccinated population. The total non-vaccinated population, is subdivided into non-vaccinated susceptible (*S*_*u*_), latent (*E*_*u*_), presymptomatic infectious (*I*_*pu*_), symptomatic infectious (*I*_*iu*_), asymptomatic infectious (*I*_*au*_), confirmed cases (*I*_*cu*_), hospitalized (*I*_*hu*_), and recovered (*R*_*u*_). The vaccinated population is subdivided into similar classes with the subscript *u* replaced with *v*. Hence, the total unvaccinated and vaccinated populations are

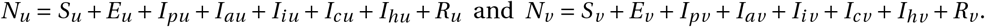

A schematic depiction of the framework is given in Fig. 2, while the variables and parameters are further described in Tables S3-S4 of the supplementary Information (SI).

**Fig. 2:**
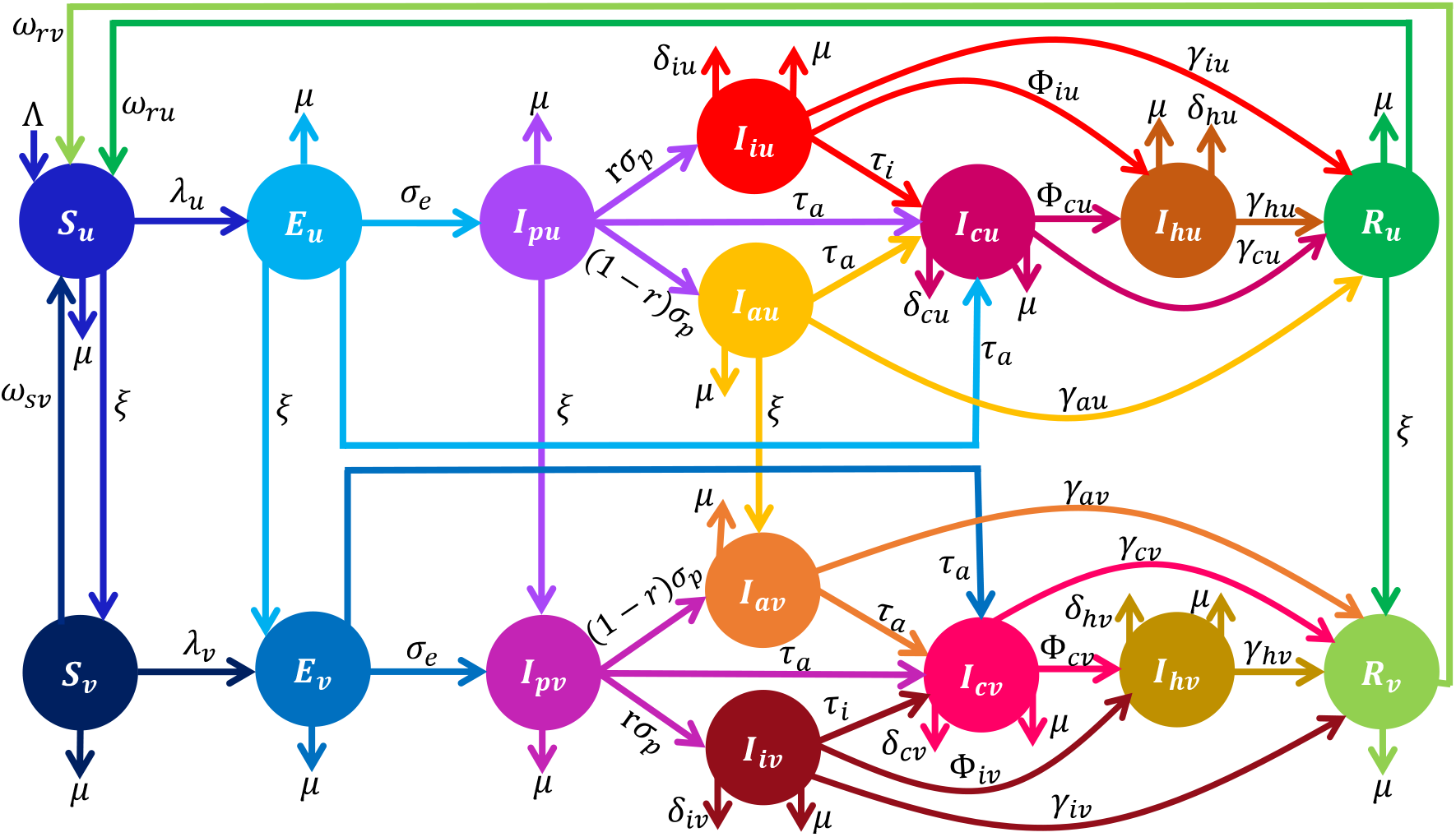
Schematic diagram of the vaccine-structured model depicting the flow of individuals between compartmentalized classes (based on disease and vaccination status). The total population (*N*) consist of unvaccinated (*N*_*u*_) and vaccinated (*N*_*v*_) groups. The unvaccinated population is made of susceptible (*S*_*u*_), latent (*E*_*u*_), presymptomatic infectious (*I*_*pu*_), symptomatic infectious (*I*_*iu*_), asymptomatic infectious (*I*_*cu*_), confirmed infectious (*I*_*cu*_), hospitalized (*I*_*hu*_), and recovered (*R*_*u*_) classes. Also, the vaccinated population is made of similar classes with the subscript *u* replaced with *v*. Descriptions of the model variables and parameters (rates) are provided in Tables S3-S4 of the SI.

In addition to the parameters described in Section 2.1, unvaccinated latent, presymptomatic infectious, asymptomatic infectious, and recovered individuals are vaccinated at per capita rate *ξ*. As a result of therapeutic benefits of COVID-19 vaccines in reducing severe infection, hospitalization, and mortality, the transmission rates, hospitalization, disease-induced mortality, and recovery rates for the non-vaccinated and vaccinated populations are different and distinguished through the use of the additional subscripts *u* and *v*, respectively. Additionally, natural (vaccine-derived) immunity for unvaccinated (vaccinated) recovered individuals wanes at per capita rate *ω*_*r u*_ (*ω*_*r v*_). The brief description above, together with the schematics (Fig. 2) and the variable and parameter descriptions in Tables S3-S4 of the SI lead to the following system of ordinary differential equations for the population dynamics of non-vaccinated individuals:

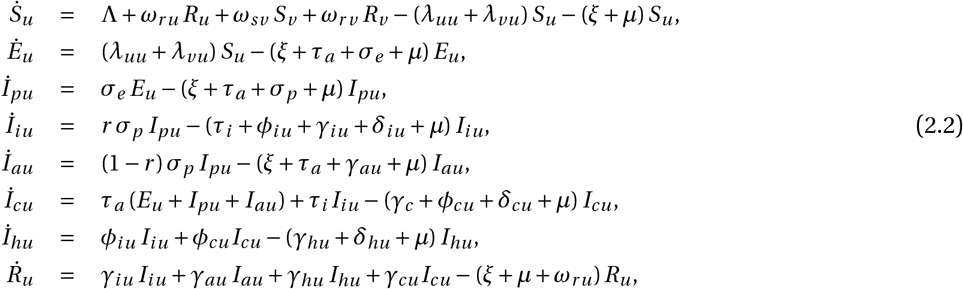

Also, using the brief description above, together with the schematics in Fig. 2 and the variable and parameter descriptions in Tables S3-S4 of the SI, we obtain the following system of equations for the dynamics of vaccinated individuals:

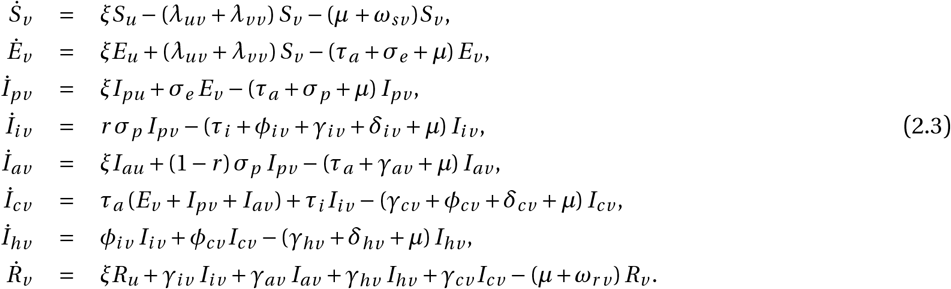

The vaccine-structured model is described by Eqs. (2.2)-(2.3), with the forces of infection *λ*_*mn*_, *m, n* ∈ {*u, v* } (depicting transmission from a *jm, j* ∈ {*p, a, i, c, h*} infectious class to an *n* susceptible class) are given by

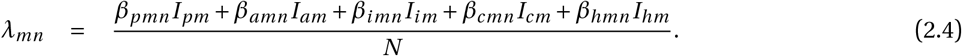

### 2.3. Model 3: Integrated (hybrid) vaccine-structured and age-structured model

The vaccine-structured model described by Eqs. (2.2)-(2.3) is now extended to account for age-structure. Here, only two age-groups based on COVID-19 risk are considered. Group 1 denoted by the additional subscript 1 consists of individuals below 65 years old (youths), while Group 2 denoted by the additional subscript 2, consists of individuals aged 65 and above (elderly). It should be mentioned that in addition to being at high risk (due to underlying health conditions and weakened immune systems) individuals aged 65 and above, have significantly different contact and mixing behaviors compared to individuals below 65 [13, 31, 35]. Other studies on COVID-19 have also broken the population into two-age groups [11, 23, 32, 33]. It should be noted that although, we have considered only two age groups here, more than two age groups defined based on disease risk, possibility of mixing, severity of disease, etc., can be accounted for. Based on our two age-group assumption, we have four coupled systems of equations that characterize the population dynamics of 1) unvaccinated individuals in age Group 1 (Eqs. (2.5)), 2) vaccinated individuals in age Group 1 (Eqs. (2.7)), 3) unvaccinated individuals in age Group 2 (Eqs. (2.7)), and 4) vaccinated individuals in age Group 2 (Eqs. (2.8)). Group 1 individuals progress to Group 2 at per capita rate, *ρ*. The total unvaccinated Group 1 population (*N*_*u*1_), vaccinated Group 1 population (*N*_*v* 1_), unvaccinated Group 2 population (*N*_*u*2_), and vaccinated Group 2 population (*N*_*v* 2_) are:

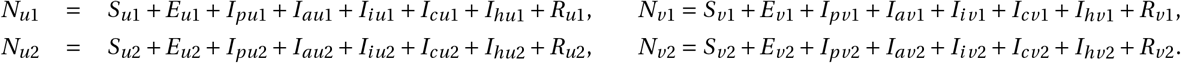

Schematics of the integrated or hybrid vaccine-structured and age-structured model are depicted in Fig. 3, the model equations are given in Eqs. (2.5)-(2.8) and the forces of infection are given by Eq. (2.9).

**Fig. 3:**
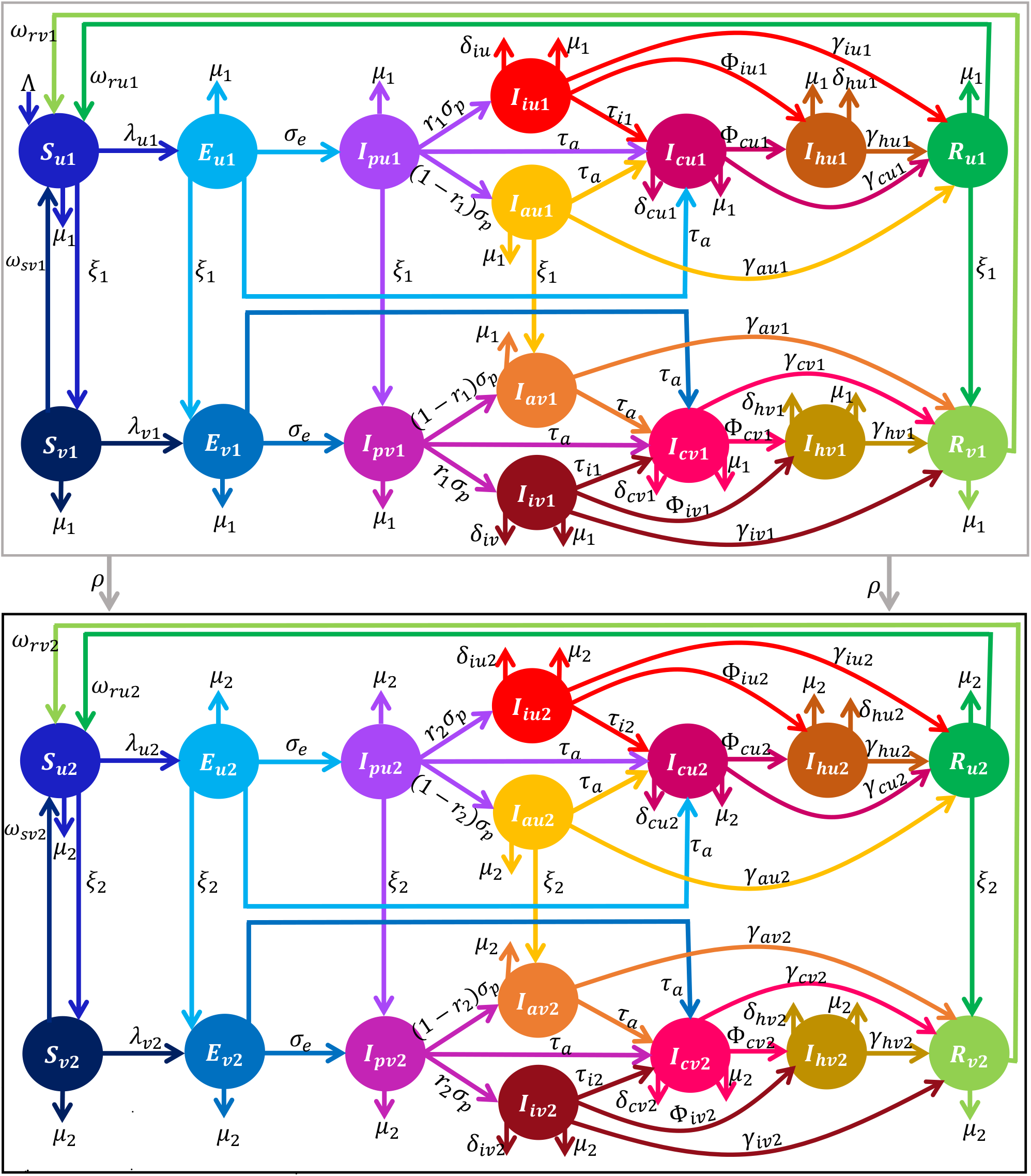
Schematics of the integrated vaccine-structured and age-structured model depicting the flow of individuals between compartmentalized classes (based on age, vaccination, and disease status). The total population (*N*) is divided into age group 1 (denoted by the subscript 1) and age group 2 (denoted by the subscript 2). Each age group is further split into the subgroups of unvaccinated (denoted by the subscripts *u*) and vaccinated (denoted by the subscript *v*), so that unvaccinated individuals in Group 1 (Group 2) carry the subscript, *u*1 (*u*2), and vaccinated individuals in Group 1 (Group 2) carry the subscript, *v* 1 (*v* 2). Furthermore, individuals in each of these subgroups are partitioned into susceptible (*S*_*mk*_), latent (*E*_*mk*_), presymptomatic infectious (*I*_*pmk*_), symptomatic infectious (*I*_*imk*_), asymptomatic infectious (*I*_*amk*_), confirmed infectious (*I*_*cmk*_), hospitalized (*I*_*hmk*_), and recovered (*R*_*mk*_), where *m* ∈ *u, v* and *k* ∈ {1, 2} classes. Group 1 individuals progress to Group 2 at per capita rate, *ρ*. Descriptions of the model variables and the other parameters (rates) are provided in Tables S5-S6 of the SI.

The population dynamics of unvaccinated individuals in age Group 1 are described by the system of equations:

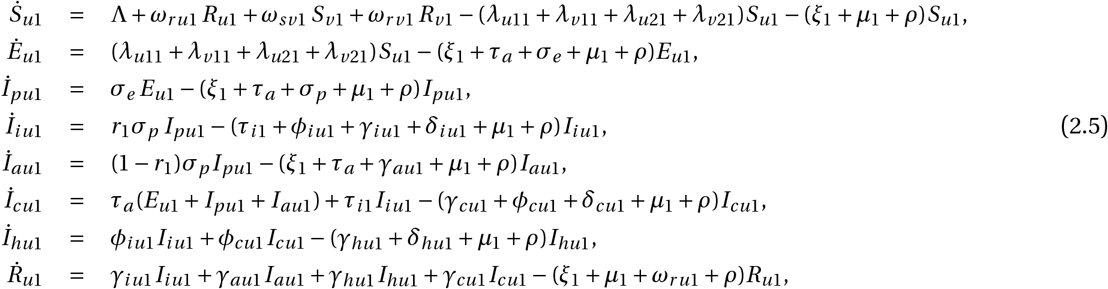

while the population dynamics of vaccinated individuals in age Group 1 are described by the system of equations:

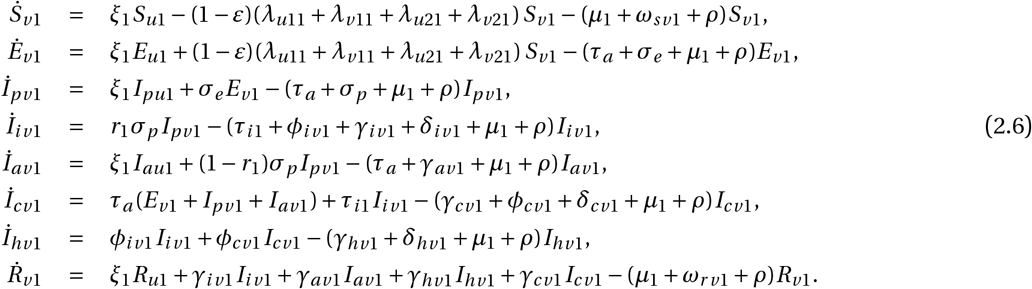

The population dynamics of unvaccinated individuals in age Group 2 are governed by the system of equations:

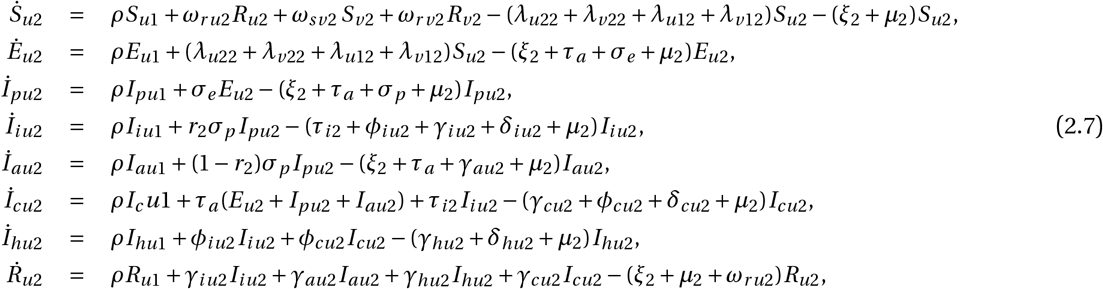

while the population dynamics of vaccinated individuals in age Group 2 are governed by the system of equations:

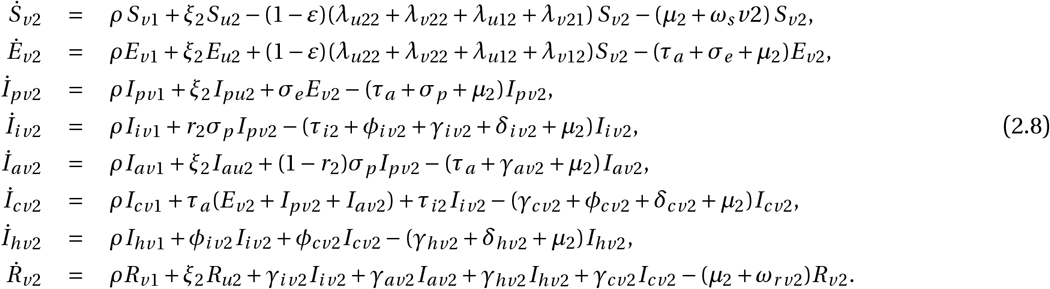

The full vaccine-age-structured model (Model 3) is described by Eqs. (2.5)-(2.8) with the forces of infection:

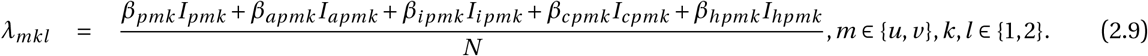

## 3. Results

## 3.1. Analytical results for the basic model

First, we demonstrate that the basic model is well-posed from a mathematical and epidemiological stand point. Consider the model (2.1) for positive time (i.e., *t* > 0) with the initial condition

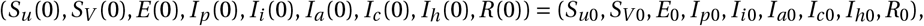

and the epidemiologically feasible region

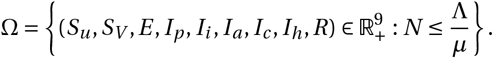

The dynamics of the total population (*N*) is described by the equation

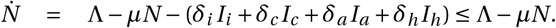

This leads to *N* (*t*) ≤ *N*_0_*e*^−*µt*^ + Λ 1 − *e*^−*µt*^ /*µ*, where *N* (0) = *N*_0_ is the initial total population size. Hence, *N* (*t*) → Λ/*µ* as *t* → ∞. Accordingly, all solutions of the basic model (2.1) that originate from Ω remain trapped in Ω. That is, Ω is positively-invariant and attracting for the model (2.1).

The disease-free equilibrium of Eqs. (2.1) is

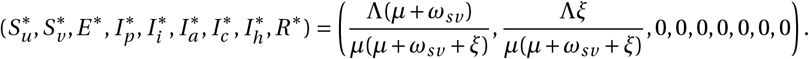

Through the next generation matrix method [36, 37], we obtain the following basic reproduction number for Model 1 (Eqs. (2.1)) in the absence of vaccination or any control measure (see Section 2.1 of the SI for details of the calculations):

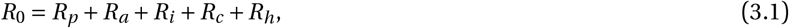

where

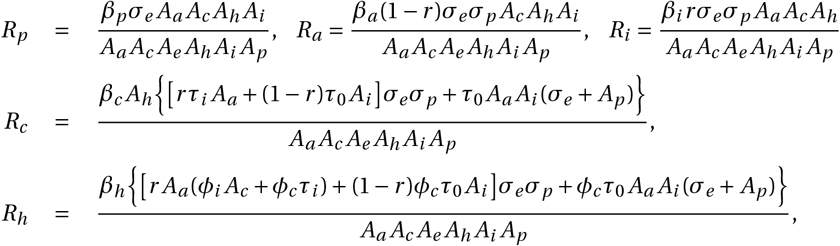

*A*_*a*_ = *τ*_0_ +*γ*_*a*_ +*μ, A*_*c*_ = *γ*_*c*_ +*ϕ*_*c*_ +*δ*_*c*_ +*μ, A*_*e*_ = *τ*_0_ +*σ*_*e*_ +*μ, A*_*h*_ = *γ*_*h*_ +*δ*_*h*_ +*μ*, and *A*_*i*_ = *τ*_*i*_ +*ϕ*_*i*_ +*γ*_*i*_ +*δ*_*i*_ +*μ, A*_*p*_ = *τ*_0_ +*σ*_*p*_ +*μ*. In the presence of vaccination and/or other control and mitigation measures, the control reproduction number is

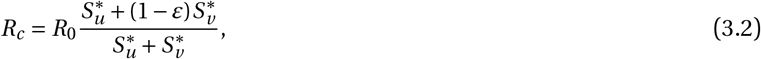

where Λ(*µ* + *ω*_*sv*_)/[*µ*(*µ* + *ω*_*sv*_ + *ξ*)] and Λ*ξ*/[*µ*(*µ* + *ω*_*sv*_ + *ξ*)]. Let *f*_*v*_ be the proportion of the susceptible population to be vaccinated to attain vaccine-derived herd immunity, then from the equation *R*_*c*_ = 1, we obtain the herd-immunity threshold

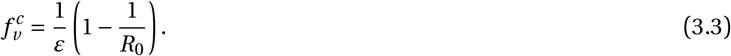

This, together with the methodology for calculating the reproduction number leads to the following result on the local stability of the disease-free equilibrium:

### Theorem 3.1.

*Achieving vaccine-derived herd immunity (and hence eliminating COVID-19) in WA using an imperfect anti-COVID-19 vaccine is possible if* 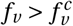 *(i*.*e*., *if R*_*c*_ < 1*) and impossible if* 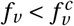 *(i*.*e*., *if R*_*c*_ > 1).

This analysis can be extended to Models 2 and 3 with slight modifications (see Section 3 of the SI).

### 3.2. Model fitting and parameter estimation

Some of the parameters for Models 1-3 (i.e., Eqs. (2.1), (2.2)-(2.3), and (2.7)-(2.8)) are available in the literature, while others are unknown. In this section, the unknown parameters for each of the three models are estimated by fitting the corresponding model to daily confirmed COVID-19 case data for the sixteen West African countries (Benin, Burkina Faso, Cape Verde, Côte D’Ivoire, Gambia, Ghana, Guinea, Guinea-Bissau, Liberia, Mali, Mauritania, Niger, Nigeria, Senegal, Sierra Leone, and Togo) [9, 38]. To incorporate the entire COVID-19 data from the onset of the pandemic in WA (i.e., from February 28, 2020) to May 24, 2022, a simplified version of the basic model (2.1) with no vaccination is fitted to data for the pre-vaccination period (i.e., the period from February 28, 2020 to May 30, 2021), while each of the three models (i.e., Eqs. (2.1), (2.2)-(2.3), and (2.7)-(2.8)) is fitted to the data for the vaccination period. Since data for WA is reported by country and not for the entire region, the data used for fitting the models is obtained by aggregating the daily countrywise confirmed COVID-19 case data. Using raw (daily case) data to fit the models is important in avoiding common errors that might arise with cumulative case data [39]. The model fitting and parameter estimation was performed using a nonlinear least-squares algorithm implemented in MATLAB (version R2021a) that minimizes the sum of the squared differences between the number of daily confirmed cases from the data (for the period from February 28, 2020 to May 24, 2022 and the number of daily confirmed cases from each of the models for the same period. Furthermore, a bootstrapping approach [40] with 1, 000 replicates was used to compute the 95% confidence intervals for the fitted parameters, while the rest of the confirmed daily case data from March 15, to May 24, 2022, and cumulative case data for the entire period from February 28, 2020 to May 24, 2022 were used for validation purposes. The fits for Models 1-3 (i.e., Eqs. (2.1), (2.2)-(2.3), and (2.7)-(2.8)) are illustrated in Fig. 4 (a)-(c), respectively, while an illustration of how well each of the models matches the confirmed case data is illustrated in Fig. 4 (d)-(f). In each of the figures for the confirmed new daily cases, the green curve depicts the model validation using the portion of the data that was not used for fitting. The estimated parameters and their 95% confidence intervals, together with the corresponding baseline values of the control reproduction numbers of the models are tabulated in the online supplementary information (Tables S7-S11).

**Fig. 4:**
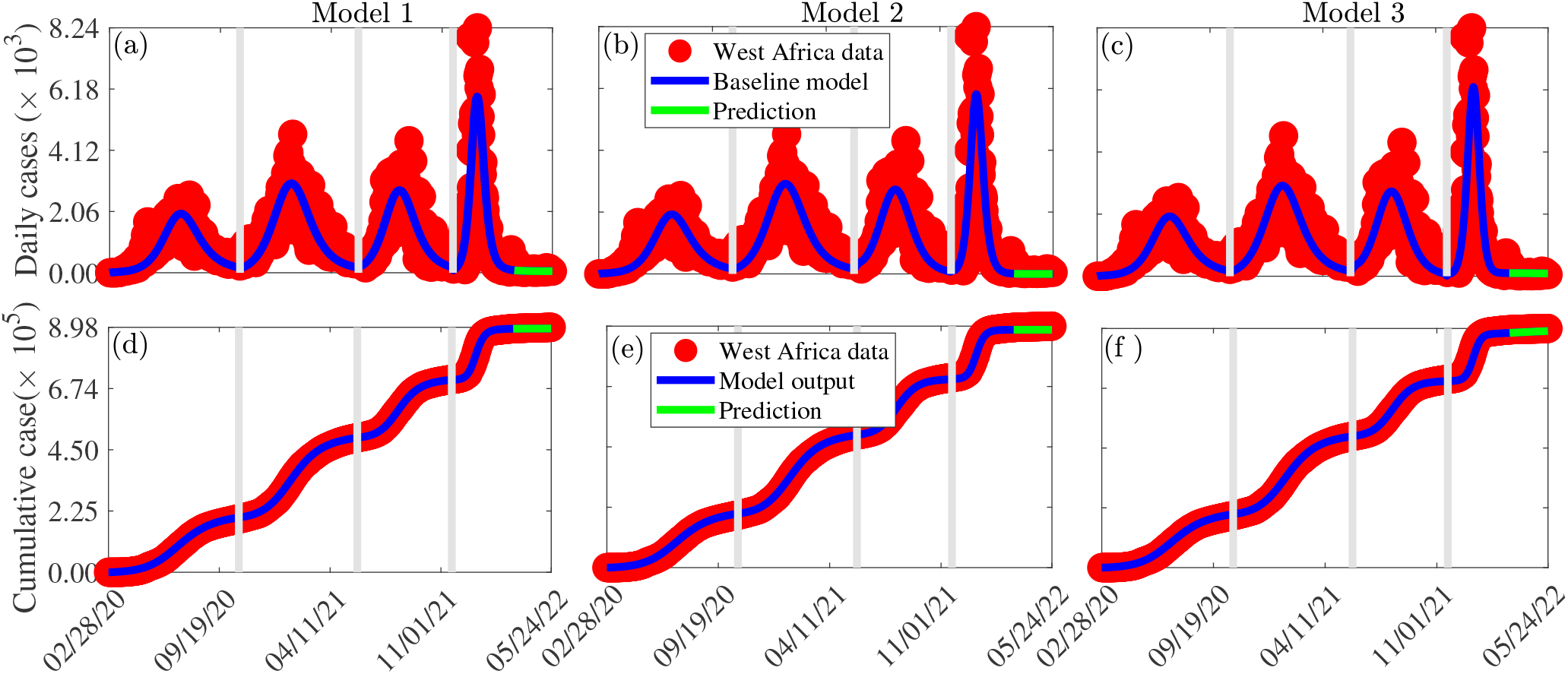
(a)-(c): Time series plots depicting the fitting of Models 1-3 (i.e., Eqs. (2.1), (2.2)-(2.3), and (2.7)-(2.8)) to confirmed daily case data for WA from February 28, 2020 to May 24, 2022. (d)-(f): Simulations of Models 1-3 using the estimated model parameters showing how well the cumulative number of confirmed COVID-19 cases from the model matches those from the West African data. The green curve, i.e., the portion of the curve from March 15, 2022 to May 24, 2022 demonstrates the performance of the models in predicting daily and cumulative COVID-19 cases in WA. The fixed and fitted parameter values are given in Tables S7-S11 of the (SI).

### 3.3. Global sensitivity analysis

Since uncertainty or variability in some model parameters can lead to substantial uncertainty or variability in some model outputs (response functions), it is instructive to carry out an uncertainty and sensitivity analysis to identify model parameters that significantly influence the model response functions. Here, the Latin Hypercube sampling (LHS) and Partial Rank Correlation Coefficient (PRCC) methodology, as well as the associated MATLAB code provided by [41] are used to perform a global uncertainty and sensitivity analysis. Assuming a uniform distribution between the minimum and maximum values of the parameters, the range of each parameter is split into 5, 000 equal sub-intervals. A Latin-Hypercube Sampling matrix, which is used to solve each model system, is generated by sampling at random without replacement from each parameter distribution. The simulation results are used to compute PRCCs corresponding to the response function (the peak number of confirmed daily COVID-19 cases in WA). The results of this analysis, depicted in Fig. 5, show that uncertainty or variability in the community transmission rate of asymptomatic infectious individuals (*β*_*a*_ for Model 1, *β*_*au*_ for Model 2, and *β*_*au*1_ for Model 3), the recovery rates of asymptomatic infectious individuals (*γ*_*h*_, *γ*_*au*_, and *γ*_*au*1_), and the detection rates of symptomatic infectious individuals (*τ, τ*_*iu*_, and *τ*_*iu*1_), will generate the greatest uncertainty or variability in the peak number of COVID-19 cases in WA. Also, changes in the transmission rates of presymptomatic infectious individuals (*β*_*p*_, *β*_*pu*_, and *β*_*pu*1_), the recovery rates of symptomatic infectious individuals (*γ*_*i*_, *γ*_*iu*_, and *γ*_*iu*1_), the proportion of pre-symptomatic infectious individuals who develop COVID-19 symptoms at the end of the incubation period (*r* for Models 1 and 2 and *r*_1_ for Model 3) and the hospitalization rates of symptomatic infectious individuals (*ϕ*_*i*_, *ϕ*_*iu*_, and *ϕ*_*iu*1_), are amongst the most influential parameters with significant impact on the response function. This analysis shows that asymptomatic infectious individuals, especially those in from age Group 1 (youths) are among the leading drivers of the pandemic in WA and that reducing disease transmission among asymptomatic infectious individuals (mostly the youths) will go a long way in curtailing the burden of the pandemic.

**Fig. 5:**
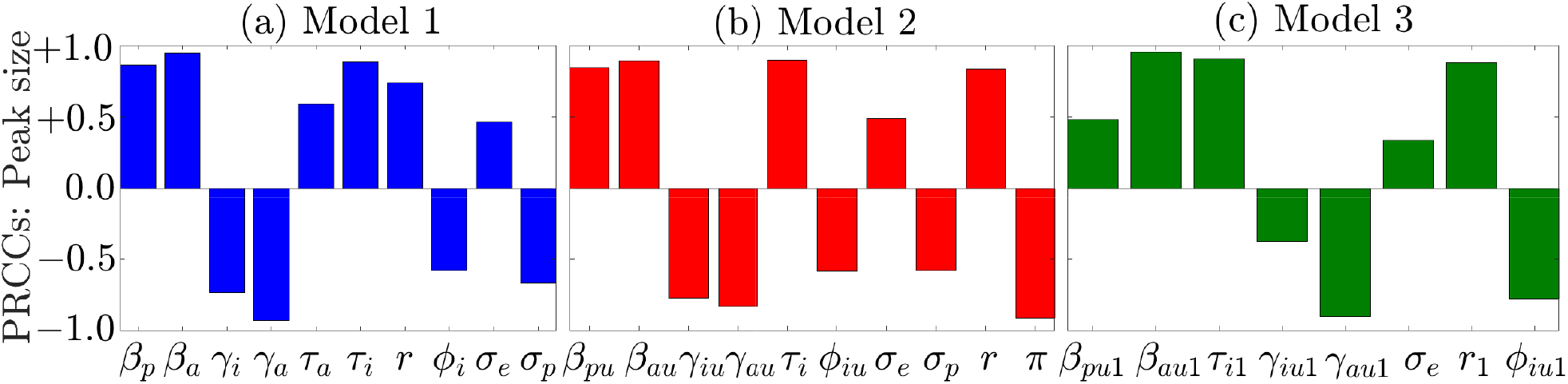
Partial Rank Correlation Coefficients (PRCCs) depicting the influence of selected model parameters on the peak number of new daily COVID-19 cases during the first Omicron wave (i.e., the overall 4^*th*^ wave) of the COVID-19 pandemic in WA for (a) Model 1, (b) Model 2, and (c) Model 3. Only parameters for which the magnitude of the PRCCs are greater than or equal to 0.3 are displayed. The baseline parameters values used for this simulation are given in Tables S7-S11.

### 3.4. Numerical simulation results

In this section, Models 1-3 (i.e., the models given by Eq. (2.1), Eqs. (2.2)-(2.3), and Eqs. (2.5)-(2.8)) are simulated to assess the impact of vaccination, control measures that result in a reduction in disease transmission, relaxation of control measures, and the impact of more transmissible new COVID-19 variants on the burden of the COVID-19 pandemic in WA. Unless otherwise specified, by vaccinated individuals, we are referring to fully vaccinated individuals (i.e., individuals who have received all required vaccine, including booster doses). Also, unless otherwise specified, the fixed and estimated baseline parameters in Tables S7-S11 of the SI are used for the simulations.

#### 3.4.1. Investigating the impact of vaccine efficacy and vaccine coverage on the burden of COVID-19 in WA

Contour plots of control reproduction number (*R*_*c*_) of each of the three models (i.e., Eqs. (2.1), Eqs. (2.2)-(2.3), and Eqs. (2.5)-(2.8)) plotted as functions of the efficacy of vaccines (*ε*) and the proportion of vaccinated individuals (*f*_*v*_) are presented in Fig. 6. It should be mentioned that while Model 3 offers a chance for vaccine prioritization between age-groups, Models 1-2 do not. The results of Model 1 show that to reduce the control reproduction number below unity (i.e., to contain the pandemic) in WA using only the widely available vaccines in the region (e.g., the AstraZeneka, Sinopharm, sputnik light, Covaxin, Sputnik V, Johnson and Johnson, Sinovac, etc., vaccines with an average efficacy of *ε* ≈ 0.67), at least 84% of the West African population (i.e., ≈ 357 million people) must be vaccinated (Fig.6 (a)). Lower vaccine coverages are required for Models 2-3. Specifically, with Model 2, at least 73% of the population of WA (i.e., ≈ 306 million people) must be vaccinated (Fig.6 (b)), while using Model 3, at least 68% of the West African populace (i.e., ≈ 289 million people) must be vaccinated (Fig.6 (c)) in order to reduce the control reproduction number below one. Hence, the vaccine-derived herd immunity thresholds for Models 1-3 are 84%, 73%, and 68%, respectively. If the West African populace adopts a vaccination policy that relies on the use of highly effective vaccines such as the Pfizer or Moderna vaccines with efficacies of ≈ 95% [42, 43], these thresholds reduce to 60% for Model 1, 51% for Model 2, and 48% for Model 3. If on the other hand, vaccines with lower efficacies are adopted, then containing the pandemic in WA will become more difficult. In particular, if only the vaccine with the lowest efficacy of ≈ 51% is used in the sub-region, Model 1 indicates that reducing the control reproduction number before one will be impossible even if the entire West African population is vaccinated (Fig.6 (a)). With Model 2, at least 95% of the population must be vaccinated to reduce the control reproduction number below one (Fig.6 (b)), while with Model 3, at least 90% vaccine coverage is required to contain the disease (Fig.6 (c)). Additional coverage thresholds for corresponding vaccine efficacies are given in Table S14 of SI. To summarize, prioritizing highly effective vaccines is important in curtailing the burden of the COVID-19 pandemic in WA. Also, not accounting for age-structure overestimates the effort required to contain COVID-19 in WA.

**Fig. 6:**
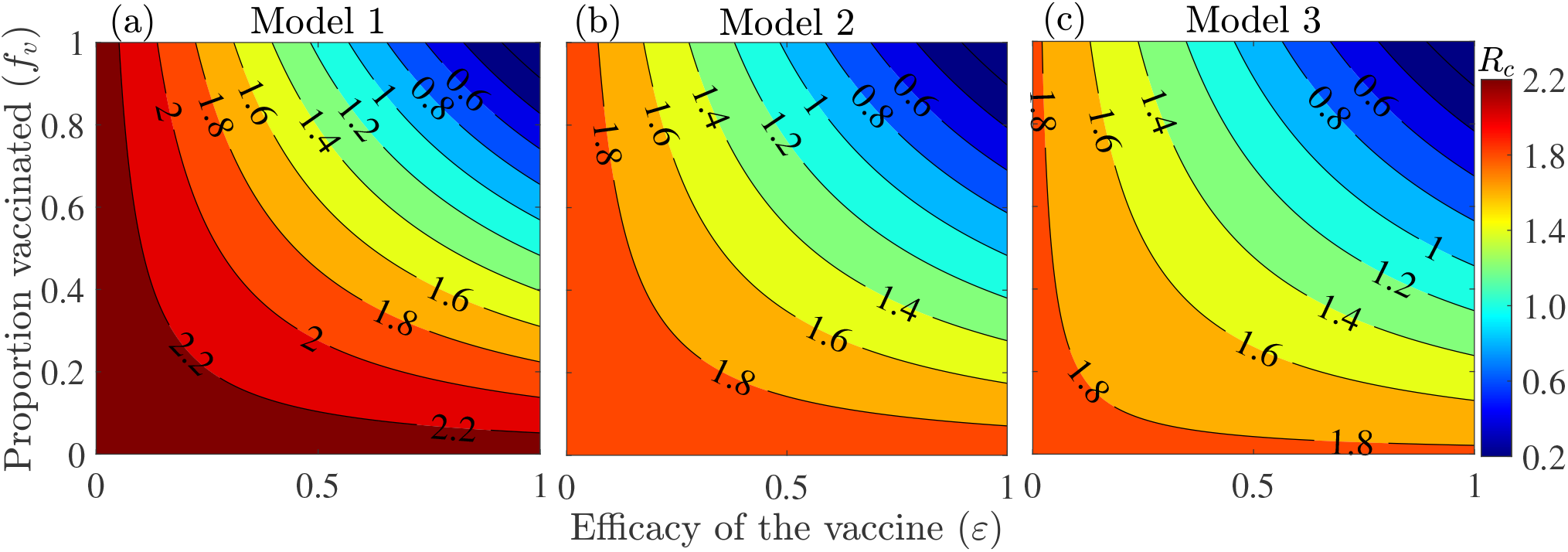
Contour plots of the control reproduction number (*R*_*c*_) of (a) Model 1 (Eq. (2.1)), (b) Model 2 (Eqs. (2.2)-(2.3)), and (c) Model 3 (Eqs. (2.5)-(2.8)) as functions of vaccine coverage (*f*_*v*_) and the protective vaccine efficacy against SARS-CoV-2 (*ε*). The other parameter values used for the simulations are provided in Tables S7-S11 of the SI.

#### 3.4.2. Investigating the impact of vaccinating youths versus the elderly on the burden of COVID-19 in WA

Figure 7 depicts contour plots of the control reproduction number (*R*_*c*_) of Model 3 as functions of the vaccination rate of the youths (*ξ*_1_) and the vaccination rate of the elderly (*ξ*_2_) to assess the combined impact of vaccine prioritization by age-group and additional mask-use compliance. The results show that reducing the control reproduction number below one by vaccinating only youths or the elderly is unattainable, even when the vaccination policy is complemented with up to a 20% increase in mask-use compliance, unless when a very high number of West Africans are vaccinated daily. However, the disease can be contained if both youths and the elderly are vaccinated at specific target rates. In particular, for the case with no additional transmission reducing measure from baseline, 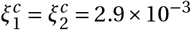 is identified as a threshold vaccination rate for reducing the control reproduction number below one (Fig.7 (a)). This corresponds to vaccinating 1.1 million youths and 0.2 million elderly daily. To reduce the control reproduction number below one, the vaccination rate of the elderly must be greater than that for youths when 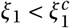, and the vaccination rate of the elderly must be less than that for youths when 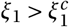. This threshold vaccinating rate will be used as a point of reference to assess the impact of relative changes in the vaccination rates of youths and the elderly. Specifically, if the threshold number of youths vaccinated is reduced by 10% (i.e., if the vaccination rate of youths is reduced from *ξ*_1_ = 2.9 × 10^−3^ to *ξ*_1_ = 2.6 × 10^−3^), then at least 24% more elderly individuals must be vaccinated (i.e., the vaccination rate of the elderly must increase from *ξ*_2_ = 2.9 × 10^−3^ to *ξ*_2_ = 3.6 × 10^−3^) to reduce the reproduction number below one. Additionally, if the threshold number of youths vaccinated is increased by 10% (i.e., if the vaccination rate of youths is now *ξ*_1_ = 3.2 × 10^−3^), then 14% less elderly people are required to be vaccinated (i.e., *ξ*_2_ = 2.5×10^−3^) to reduce the reproduction number below one. On the other hand, if the threshold number of the elderly people vaccinated is reduced by 10% (i.e., if *ξ*_2_ = 2.6×10^−3^), then at least 7% more youths must be vaccinated (i.e., *ξ*_1_ = 3.1×10^−3^) to reduce *R*_*c*_ below one. Furthermore, if the threshold number of the elderly vaccinated is increased by 10% (i.e., if *ξ*_2_ = 3.2 × 10^−3^), then 3% less elderly are required to be vaccinated (i.e., *ξ*_1_ = 2.8 × 10^−3^) to reduce *R*_*c*_ below one.

**Fig. 7:**
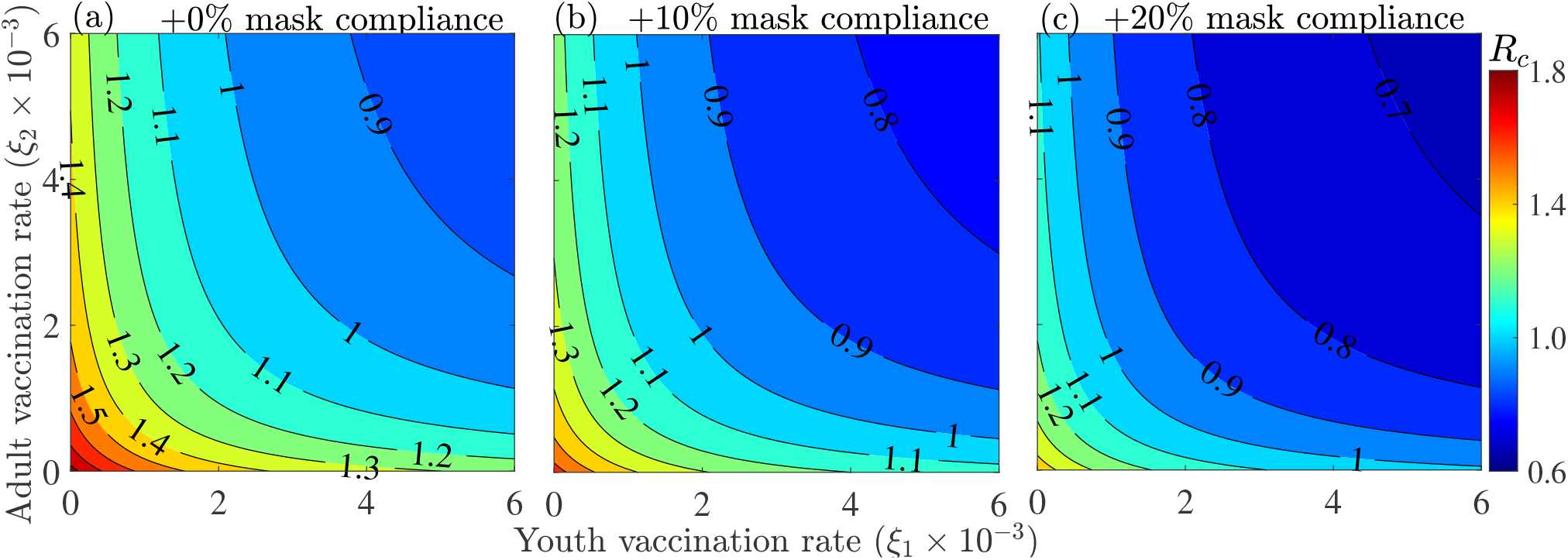
Contour plots of the control reproduction number (*R*_*c*_) of Model 3 (Eqs. (2.5)-(2.8)) as functions of the vaccination rates of youths (*ξ*_1_) and the elderly (*ξ*_2_) for different levels of additional mask-use compliance (*c*_*m*_): (a) no additional mask-use compliance, (b) +10% mask compliance, and (c) +20% mask compliance. The other parameter values used for the simulations are provided in Tables S7 & S11 of the SI.

For the case in which additional transmission rate reducing controls are implemented, the numbers of youths and elderly required to be vaccinated to bring the control reproduction number below one are reduced. In particular, if the vaccination rate of youths is *ξ*_1_ = 2.9 × 10^−3^ *per day* and the current level of transmission reducing control measures is increased by 10%, then the vaccination rate of the elderly must be *ξ*_2_ = 1.1 × 10^−3^ *per day* (Fig.7 (b)), while if the current level of transmission reducing control measures is increased by 20%, then the vaccination rate of the elderly must be *ξ*_2_ = 0.4 × 10^−3^ to reduce the reproduction number below one (Fig.7 (c)). However, if the vaccination rate of the elderly is *ξ*_1_ = 2.9 × 10^−3^ *per day* and the current level of transmission reducing control measures is increased by 10%, then the vaccination rate of youths must be *ξ*_2_ = 1.5 × 10^−3^ *per day* (Fig.7 (b)), while if the current level of transmission reducing control measures is increased by 20%, then the vaccination rate of youths must be *ξ*_2_ = 0.7 × 10^−3^ to reduce the reproduction number below one (Fig.7 (c)). It should be noted that for the 10% transmission rate reducing scenario, a 34.5% reduction in the vaccination threshold is attained (i.e., *ξ*_1_ = *ξ*_2_ = 1.9×10^−3^.), while for the 20% transmission rate reducing scenario, there is a 58.6% reduction in the threshold vaccination rate (i.e., *ξ*_1_ = *ξ*_2_ = 1.2 × 10^−3^). To summarize, vaccinating more youths than the elderly is a good strategy for containing the COVID-19 pandemic in WA, while vaccinating more youths and the elderly simultaneously is a better strategy. Under any of these scenarios, prospects of containing the pandemic are improved if the mask-use compliance level is raised.

#### 3.4.3. Investigating the impact of vaccination on the dynamics of COVID-19 in WA

The three models given by Eqs. (2.1), Eqs. (2.2)-(2.3), and Eqs. (2.5)-(2.8) are simulated to assess the impact of vaccination on the number of confirmed daily COVID-19 cases in WA. The results obtained (Fig. 8), show that vaccination has a significant impact in reducing the peak number of confirmed daily cases in WA. Specifically, if the vaccination rate in Model 1 is maintained at its baseline value of 1.174 × 10^−5^ (i.e., only ≈ 5, 000 West Africans are vaccinated daily), there is a possibility of multiple pandemic waves, with the next wave peaking by mid November, 2022 and milder than the Omicron wave. If the vaccination rate is increased from baseline to 2.9×10^−3^ *per day* (i.e., ≈ 1.2 million West Africans are vaccinated daily), a 14% reduction in the baseline peak number of daily confirmed cases will be recorded using Model 1 (comparing the major blue and the magenta peaks in Fig. 8 (a)), while a 21% reduction in the baseline peak size will be recorded using Model 2 (comparing the blue and magenta curves in Fig. 8 (b)). In this case, elimination is possible by April 18, 2022 using Model 2, while another minor pandemic wave that will peak by mid February 2023 is possible for Model 1. For the same vaccination rate, an additional 7% reduction in the peak number of confirmed cases is recorded with Model 2 compared to Model 1 (comparing the difference between the major blue and magenta peaks in Fig. 8 (a) and the difference between the major blue and magenta peaks in Fig. 8 (b)). Furthermore, if the vaccination rate in is increased from its baseline value to 1.2 × 10^−2^ *per day* (i.e., if ≈ 2.3 million West Africans are vaccinated daily) then more significant reductions in the peak number of confirmed daily cases are recorded for both Models 1 and 2. Specifically, a 26% reduction is recorded using Model 1 (comparing the blue and green curves in Fig. 8 (a)), while a 41% reduction is recorded using Model 2 (comparing the blue and magenta curves in Fig. 8 (b)). It should be noted that an additional reduction of 25% in the peak number of confirmed cases is recorded if Model 2 is used instead of Model 1 (comparing the difference between the blue and green curves in Fig. 8 (a) and the difference between the blue and green curve in Fig. 8 (b)). Thus, Model 1 overestimates diseased burden and the required effort to contain the disease compared to Model 2.

**Fig. 8:**
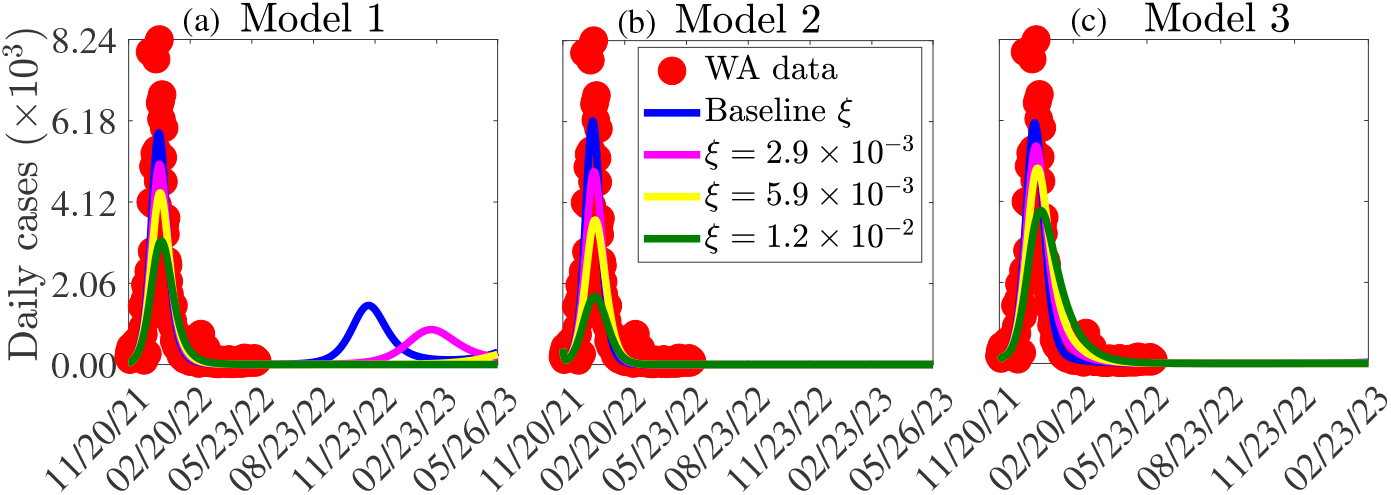
Simulations of (a) Model 1, (b) Model 2, and (c) Model 3 illustrating the number of new daily COVID-19 cases in West Africa for different vaccination rates (*ξ* for Models 1-2 and *ξ*_1_ and *ξ*_2_, where *ξ* = *ξ*_1_ = *ξ*_2_ for Model 3). The other parameter values used in the simulations are provided in Tables S7-S11 of the SI.

In the case of Model 3, if the vaccination rates of youths (*ξ*_1_) and elderly (*ξ*_2_) increase from their respective baseline values to 2.9 × 10^−3^ (i.e., if *ξ*_1_ = *ξ*_2_ *per day*, or 1.05 million youths and 185 thousands elderly are vaccinated daily), there is a 10% decrease in the peak number of daily confirmed cases (comparing the blue and magenta curves in Fig. 8 (c)). A further increase in the vaccination rates of youths and the elderly will lead to a more significant decrease in the peak number of daily confirmed cases. In particular, if *ξ*_1_ = *ξ*_2_ = 1.2 × 10^−2^, a 36% reduction in the pandemic peak size is recorded (comparing the blue and green curves in Fig. 8 (c)). On the other hand, if the vaccination rate of the elderly is maintained at its baseline value of *ξ*_2_ = 9.4 × 10^−6^, while that of youths is increased to *ξ*_1_ = 2.9 × 10^−3^ (i.e., ≈ 1.05 million youths are vaccinated daily), there is a 8% decrease in the peak number of daily confirmed cases (comparing the blue and the light green curves in Fig. S1 of the SI). But if the vaccination rate of youths is maintained at its baseline value of *ξ*_1_ = 2.3 × 10^−6^, while that of the elderly is increased to *ξ*_2_ = 2.9 × 10^−3^, there is a 2% decrease in the peak number of confirmed daily cases (comparing the blue and magenta curves in Fig. S1 of the SI). If *ξ*_1_ = 1.2 × 10^−2^ and *ξ*_2_ = 2.9 × 10^−3^, a 34% reduction in the peak size is recorded (comparing the blue and yellow curves in Fig. S1 of the SI), if *ξ*_2_ = 1.2 × 10^−2^ and *ξ*_1_ = 2.9 × 10^−3^, a 13% reduction in the peak size is recorded (comparing the blue and grey curves in Fig. S1 of the SI). Hence, increasing vaccination rate among youth leads to more reduction in the peak size (comparing the yellow and grey curves in Fig. S1 of the SI). To summarize, vaccinating more youths than the elderly or vaccinating both more youths and more elderly is critical in controlling the COVID-19 pandemic in West Africa.

#### 3.4.4. Investigating the impact of additional masking on the dynamics of COVID-19 in WA

To investigate the impact of additional control measures that result in a reduction in the effective transmission rate in each of the three models on the daily and cumulative COVID-19 cases in WA, the models are simulated using the following three (arbitrary) values of the transmission reduction-related parameters: 1) low additional reduction (*c*_*m*_ = 0.15), 2) moderate additional reduction (*c*_*m*_ = 0.30), and (c) high additional reduction (*c*_*m*_ = 0.45). The results obtained and displayed in Fig. 9 show the possibility of another wave with a lower peak size for the baseline, low, and moderate additional reduction in transmission scenarios if Model 1 is used. For example, under the low and moderate additional reduction in transmission rate scenarios, 21%, and 50% reductions in the minor peak sizes are recorded (comparing the minor blue peak with the minor magenta and yellow peaks in Fig. 9 (a)). In what follows, we focus on the major wave (i.e., the wave with the larger peak size). Figure 9 shows that significant reductions in the burden of COVID-19 in West Africa will be achieved with additional reduction in transmission, especially for the cases of Models 1 and 2. In particular, using Model 1 with the low additional mask compliance level, a 31% reduction in the baseline peak number of daily cases is recorded (comparing the major blue and magenta peaks in Fig. 9 (a)). For this same low additional mask compliance level, a 32% reduction in the baseline peak number of daily cases is recorded (comparing the major blue and magenta peaks in Fig. 9 (b)), while with Model 3, a 17% reduction in the baseline peak number of daily cases is recorded (comparing the major blue and magenta peaks in Fig. 9 (c)). Under the moderate additional reduction in transmission rate scenario, a 60% reduction from the baseline peak number of daily cases is recorded with Model 1 (comparing the major blue and yellow peaks in Fig. 9 (a)), a 66% reduction from the baseline peak numbers of daily cases is recorded with Model 2 (comparing the major blue and yellow peaks in Fig. 9 (b)), while a 36% reduction from the baseline peak numbers of daily cases is recorded with Model 3 (comparing the major blue and yellow peaks in Fig. 9 (c)). Even more significant reductions are obtained under the high additional reduction in transmission rate scenario. Specifically, an 88% reduction in the peak numbers of daily cases is obtained with Model 1 (comparing the major blue and green peaks in Fig. 9 (a)), a 94% reduction is obtained with Model 2 (comparing the major blue and green peaks in Fig. 9 (b)), and a 56% reduction is obtained with Model 3 (comparing the major blue and green peaks in Fig. 9 (c)). For each of the three reductions in transmission rate scenarios, the reduction in the peak size for Model 2 is lower than that for Model 1, while the reductions for Models 1 and 2 are lower than those for Model 3 (comparing the same curve colors in Fig. 9 (a)-(c)). Similar reductions are obtained for the cumulative number of confirmed cases (Fig. 9 (d)-(f)).

**Fig. 9:**
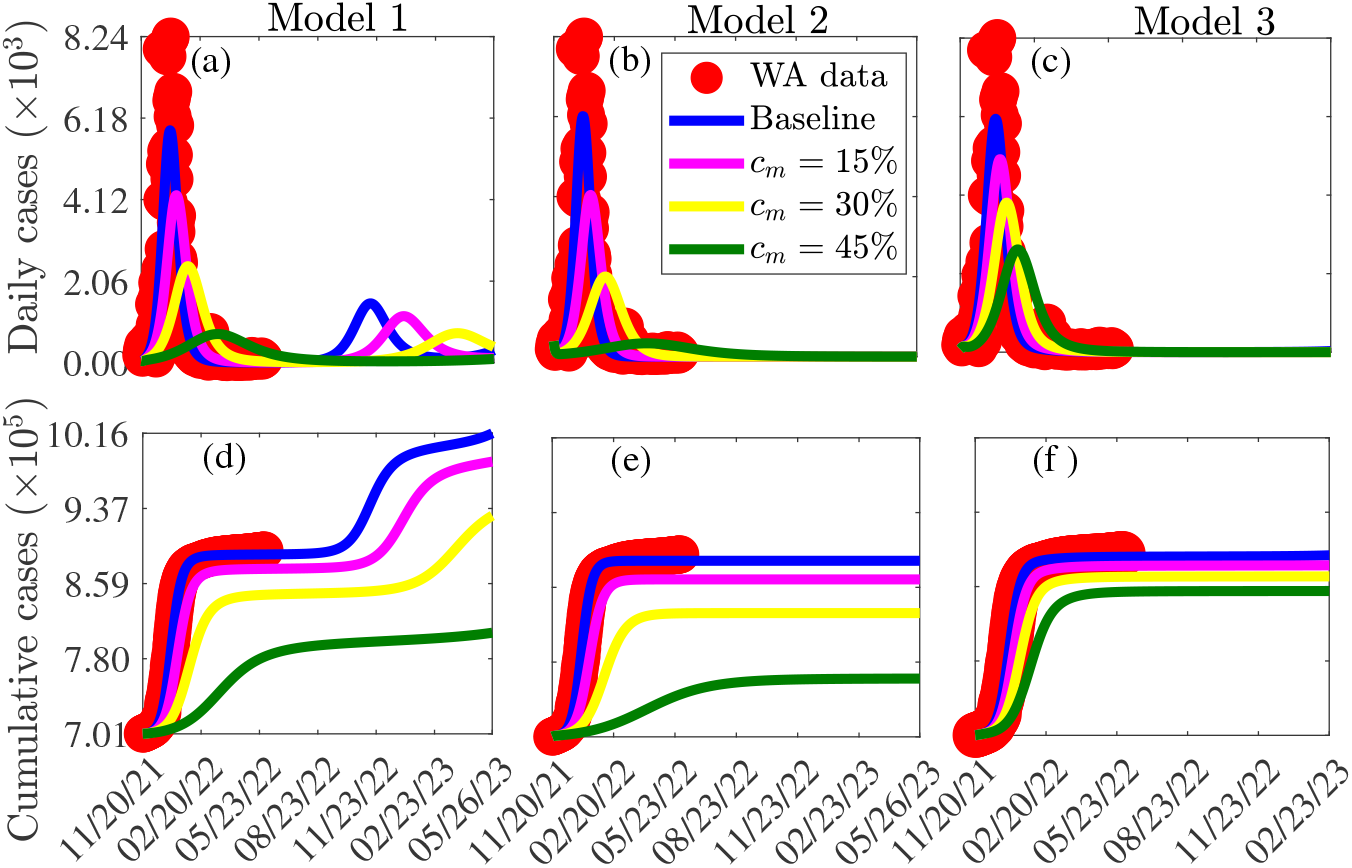
Simulations of Model 1 ((a) and (d)), Model 2 ((b) and (e)), and Model 3 ((c) and (f)) showing the daily number of confirmed cases ((a)-(c)) and the cumulative number of cases ((d)-(f)) for various additional reductions (*c*_*m*_) in the transmission rate. The values of the other parameters used for the simulations are given in Tables S7-S11 of the SI.

#### 3.4.5. Investigating the combined impact of vaccination and additional masking on the dynamics of COVID-19 in WA

The three models described by Eq. (2.1), Eqs. (2.2)-(2.3), and Eqs. (2.5)-(2.8) are simulated to explore the combined impact of vaccination and additional reduction in disease transmission (resulting from additional mask-use compliance) on the number of daily cases of COVID-19 in West Africa. Here, only the arbitrarily selected low and moderate levels, corresponding to +15% and 30% additional reductions in transmission rates are considered. The results of the simulations presented in Fig. 10 show drastic reductions in the peak number of daily cases when additional reduction in transmission is combined with increased vaccination. In particular, for a 15% additional reduction in transmission, if the vaccination rate is *ξ* = *ξ*_1_ = *ξ*_2_ = 2.9 × 10^−3^, a 42% reduction in the peak number of daily cases will be registered for Model 1 (comparing the major blue and magenta peaks in Fig. 10 (a)), a 55% reduction in the peak number of daily cases will be registered for Model 2 (comparing the major blue and magenta peaks in Fig. 10 (b)), and a 27% reduction in the peak number of daily cases will be registered for Model 3 (comparing the major blue and magenta peaks in Fig. 10 (c)).

**Fig. 10:**
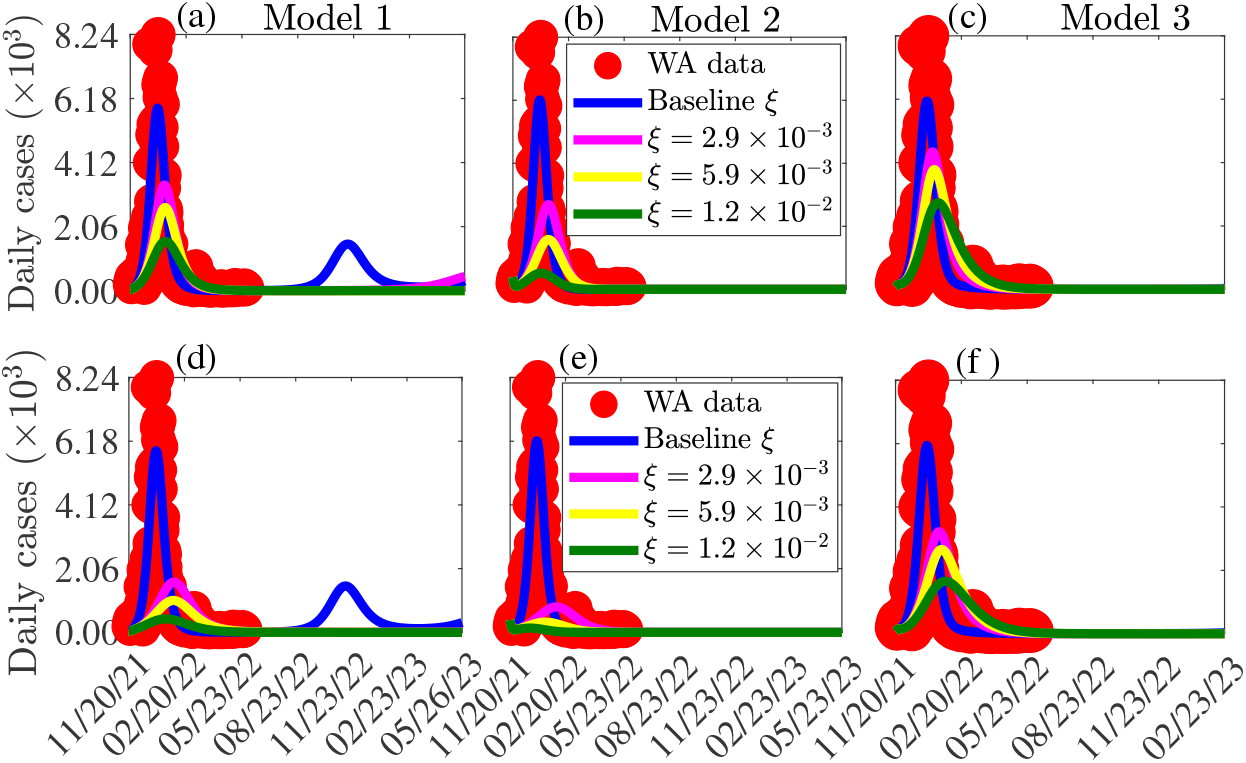
Simulations of Model 1 ((a) and (d)), Model 2 ((b) and (e)), and Model 3 ((c) and (f)) to assess the impact of a 15% additional reduction in COVID-19 transmission ((a) and (c)) and a 30% additional reduction in COVID-19 transmission ((d) and (f)) for different vaccination rates (*ξ* = *ξ*_1_ = *ξ*_2_) on the number of new daily COVID-19 cases in WA. The values of the other parameters used for the simulation are as given in Tables S7-S11 of the SI.

On the other hand, for a 15% additional reduction in transmission, if the vaccination rate is *ξ* = *ξ*_1_ = *ξ*_2_ = 1.2×10^−2^, a 73% reduction in the peak number of daily cases will be registered for Model 1 (comparing the major blue and green peaks in Fig. 10 (a)), a 91% reduction in the peak number of daily cases will be registered for Model 2 (comparing the major blue and green peaks in Fig. 10 (b)), and a 54% reduction in the peak number of daily cases will be registered for Model 3 (comparing the major blue and green peaks in Fig. 10 (c)). The reductions are even more drastic for the moderate reduction in transmission case, i.e., when *c*_*m*_ = 0.30 (Fig. 10 (d)-(f)).

### 3.5. Assessing the impact of relaxing transmission reduction measures or more transmissible new variants of COVID-19

At the time of this writing, the number of new daily cases of COVID-19 in WA was very low (≈ 100 cases *per day*). This could be attributed to limited testing, under-reporting, or the lack of reporting from some countries. The low case numbers have led to the relaxation of most of the control measures. In this section, Models 1-3 are simulated to assess the impact of reinforcing or relaxing existing transmission control strategies, as well as the emergence of new more transmissible variants of SARS-CoV-2 in WA. The results obtained (Fig. 11) show that reinforcing existing transmission control strategies can prevent subsequent waves of the pandemic from occurring, while relaxing the existing control measures can lead to additional waves in West Africa. If control measures are maintained at their current level, another wave of the pandemic with a peak size 74% less than that of the Omicron wave for Model 1 will be possible (Fig. 11 (a)). If additional measures that can result in a 50% reduction in COVID-19 transmission in WA are implemented, then the disease can be contained for each of the three models with no subsequent waves (magenta curves in Fig. 11 (a)-(c)). However, if the existing control measures are relaxed to a level that triggers a 50% increase in the effective transmission rate increases or if is a new variant that is 1.5 times more transmissible emerges, then containing the pandemic in the WA will be more difficult since this will lead to subsequent waves with peak sizes lower than those of the Omicron wave for Models 1 and 2 (black curves in Fig. 11 (a)-(b)). In this case, the size of the new wave will be 90% that of the Omicron wave for Model 1 and 69% that of the Omicron wave for Model 2. Further relaxing of the control measures will lead to more devastating subsequent waves (yellow curves in Fig. 11 (a)-(c)).

**Fig. 11:**
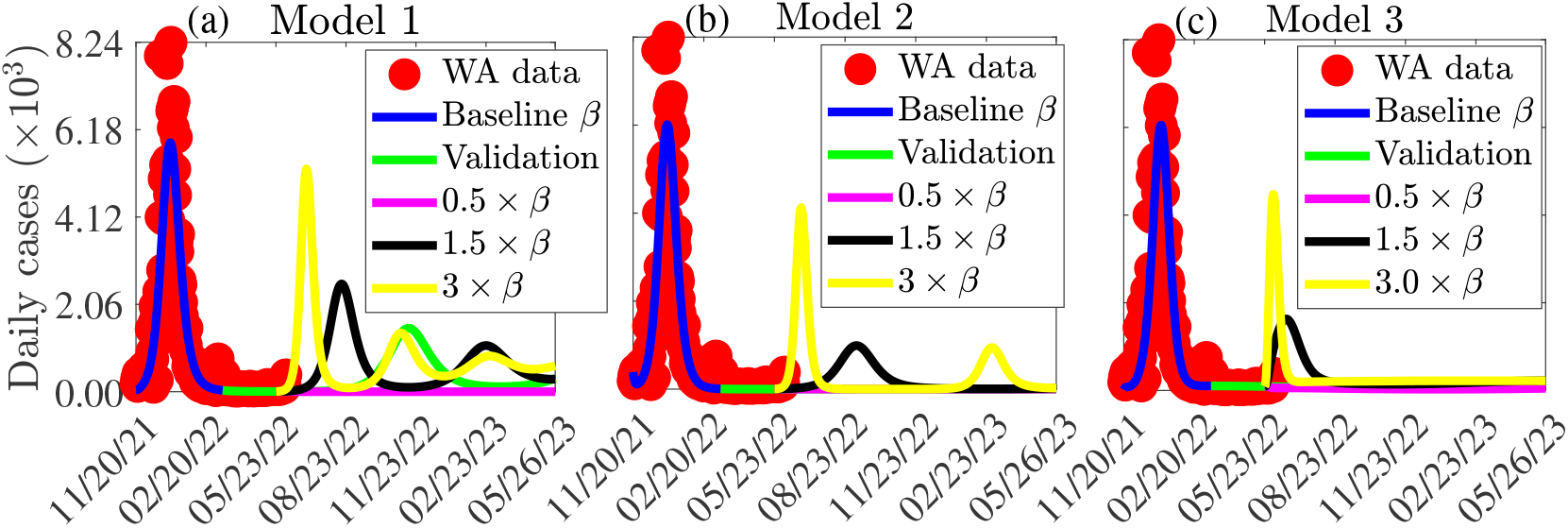
Simulations of (a) Model 1, (b) Model 2, and (c) Model 3 showing the effects of reinforcing or relaxing control measures that result in a change in the effective transmission rate or the emergence of new more transmissible variants of SARS-CoV-2 in WA. The values of the other parameters are given in Tables S7-S11.

## 4. Discussion, limitations, and conclusion

### 4.1. Discussion and limitations

A significant impact of COVID-19 has been felt in almost every nation throughout the world. Although NPIs and the design and deployment of several safe and effective vaccines for use within a very short time helped significantly in limiting the spread of the virus, COVID-19 is becoming increasingly difficult to combat due to the emergence of VOC against which existing vaccines provide only some degree of cross-protection. Additionally, limited supply of these vaccines to some regions, e.g., WA calls for prioritization of vaccine administration to certain target groups. In this study, three mathematical models that account for vaccination strategies are formulated and used to explain and predict the burden of COVID-19 in WA. In the first model, only susceptible individuals are vaccinated, while in the second model, exposed, presymptomatic, and asymptomatic individuals are also vaccinated since we are not aware of their disease status unless when they are tested. In this extended model, we track two different groups of individuals (namely, vaccinated and unvaccinated individuals) in each of the classes. The third model extends the second through the introduction of a dichotomous age structure of the population. Accounting for age structure in the dynamics of COVID-19 in WA is critical for vaccine prioritization since the risk of COVID-19 is associated with age and underlying medical conditions and since there is a wide gap between the youthful and elderly populations in WA [44, 45]. The importance of accounting for age structure in models aimed at explaining the spread and control of COVID-19 have been highlighted in [31, 46]. Standard techniques from nonlinear dynamical systems and infectious disease modeling are used to acquire rigorous insights into the dynamics of the models. These include computing the basic and control reproduction numbers and using them to establish the existence and stability of disease-free equilibria and for computing vaccine-derived herd immunity thresholds (i.e., the proportion of the population to vaccinate in order to contain the disease) of the models.

Although some parameters of each of the three models are known and available in the literature, others are unknown. To estimate the unknown parameters, each of the models is fitted to confirmed new daily COVID-19 case data from West Africa. The segment of data from February 28, 2020 to March 24, 2022 is used for fitting, while the remaining segment for the period from March 25, 2022 to May 24, 2022 is used for validation. Additionally, cumulative case data for the entire region is used for validation of the model. In both cases, there is a good match between the number of confirmed cases from the data and the number of confirmed cases from the model. The estimated parameters show that the transmission rates of asymptomatic infectious individuals are greater than those for symptomatic infectious individuals. For example, during the Omicron wave, asymptomatic infectious youths were three times more infectious than symptomatic youths, whereas for the elderly, there was no significant difference in the transmission rates of the symptomatic and asymptomatic infectious. Also, disease transmission between the elderly population is lower than between the younger population, while transmission from youths to the elderly is higher than transmission from the elderly to the youths. This is consistent with results from a study by Richard *et al*. [31], which show a relatively low contact rate among individuals aged 60 years and older in Burkina-Faso. Our study points to the fact that infectious individuals who exhibit very mild or no disease symptoms, particularly those younger than 65 years old are the main drivers of the COVID-19 pandemic in WA. This is reasonable since most of the population of WA is young and younger people have wider networks of contacts and interactions. Also, because most infected youths experience only mild or no disease symptoms at all, they are not aware that they are infected with SARS-CoV-2. These results are consistent with those in [26, 47, 48]. Curtailing SARS-CoV-2 spread by asymptomatic infectious individuals (e.g., through mass testing of youths and education on safe practices) can have a significant impact in reducing the burden of the pandemic in the West African sub-region.

The numerical values of the herd immunity thresholds computed using the known and estimated values of the parameters of the three models are 84% for Model 1, 73% for Model 2, and 68% for Model 3, which are in the range of herd immunity thresholds reported in [49, 50]. Also, the African Centers for Disease Control’s COVID-19 vaccine program recommends that 70% of the population be fully vaccinated (with 30% of this target by the African Vaccine Acquisition Trust (AVAT) program and the remaining 40% by the COVID-19 Vaccines Global Access (COVAX) in Africa [2]). Despite this recommendation, a very wide gap between the required and available number of vaccine doses still exists. Specifically, only 6.7% of the 843.2 million vaccine dose target by AVAT and only about 52.2% of the required vaccine doses from the COVAX program had been received by late June 2022 [2]. To compound matters, even with this failure in vaccine delivery, not all the delivered doses have been administered as a result of factors such as vaccine hesitancy. At the current baseline vaccination rates, it will take 5.7 years to attain herd immunity with Model 1, 5 years to attain herd immunity with model 2, and 4.6 years to attain herd immunity with Model 3. However, with the availability of more vaccine doses, if the current vaccination rates for youths and the elderly in Model 3 are tripled, then herd immunity will be attainable in WA by the end of 2023. This result aligns with studies in [51] calling for a substantial ramp up in COVID-19 vaccination in WA to achieve herd immunity. Another issue with the vaccination program in WA is that many of the vaccines used in the area are of lower efficacy. Our study shows that higher proportions of the population are required to be vaccinated to achieve vaccine-derived herd immunity if the efficacies of the vaccines used in WA are low. For example, if only the vaccine with the lowest efficacy (i.e., the Sinovac-CoronaVac vaccine with an efficacy of ≈ 51%) is used, then vaccine-derived herd immunity threshold for Model 2 (Model 3) is 95% (90%). But if the region prioritizes the use of highly efficacious vaccines such as the Pfizer-BioNTech or Moderna vaccine, the vaccine-derived herd immunity thresholds are lower (51% for Model 2 and 48% for Model 3. Computations of the reproduction numbers of the three models show that the reproduction number of Model 1 is bigger than that of Model 2, which in turn is bigger than the reproduction number of Model 3. In particular, the computed reproduction number for Model 3 (i.e., *R*_*c*_ ≈ 1.3) is in the range of the average computed reproduction numbers for WA for the same period [52]. Hence, the reproduction numbers and herd immunity thresholds of the models show that disease burden and the required control efforts are overestimated in models in which age-structure is not accounted for. Thus, accounting for age-structure is critical in explaining the control and transmission dynamics of COVID-19. This is consistent with results presented in [31, 46].

A global sensitivity analysis identifies model parameters that are associated with asymptomatic infectious individuals among the parameters with the most significant impact on the peak size of the number of confirmed COVID-19 cases for each of the three models. This, again, confirms the fact that asymptomatic infectious individuals, especially those within the youthful group are at the center of COVID-19 transmission of in WA. When most individuals within this age group become infected, they either do not exhibit any clinical symptoms of COVID-19 or they exhibit only mild symptoms. Because they constitute the greater proportion of the population and have larger contact networks, they easily spread the virus unknowingly. Therefore, preventing COVID-19 spread by asymptomatic infectious individuals (e.g., through mass testing and isolation of detected infectious youths and extensive education on safe COVID-19 practices) can have a significant impact in reducing the burden of the pandemic.

Simulations of the models show that vaccination is important in reducing the spread of COVID-19, especially if vaccines of higher efficacy than most of those currently used in WA are prioritized. Also, vaccinating a high proportion of the population below 65 years compared to that 65 and above is critical for containing the pandemic, while vaccinating more youths and the elderly simultaneously is even a better control strategy. This suggests a different vaccination policy compared to that implemented in high-income countries such as the U.S., where the elderly and most vulnerable (i.e., those with pre-existing medical conditions) were first vaccinated during the initial phase of vaccine allocation and booster vaccine dose administration [53, 54]. This strategy will be unsuccessful in sub-Saharan Africa and WA in particular with a very young population. However, it should be mentioned that while prioritizing vaccination of the elderly population is critical in reducing severe illness, hospitalization, and mortality [31], directing more vaccine administration towards the youths is essential in reducing the spread of COVID-19 [32, 33]. Additional simulations of the model show that reinforcing current control and mitigating measures, especially those measures that translate to a direct reduction in the spread of COVID-19 has a significant impact in curtailing the burden of the pandemic, especially when such measures are combined with vaccination policies. On the other hand, relaxing the current measures, or the emergence of new more transmissible variants of SARS-CoV-2 can trigger more devastating waves of the pandemic.

The accuracy of our estimates is limited by the use of incidence data in building our models, which is less precise compared to wastewater data or data on severe and hospitalized cases. These additional data sources were not readily available at the time this research was carried out. Additionally, under reporting of cases and low testing capacity within the sub-region could have affected our findings. The gap in the literature regarding the extent of social contacts among various age groups in WA inspired our use of a dichotomous age structure instead of deploying a contact matrix in assessing the impact of age structure on the transmission dynamics of COVID-19 in the sub-region. The proposed framework can be extended to account for multiple dose vaccines explicitly. The models assume that vaccine-derived natural immunity wanes completely so that individuals from the vaccinated susceptible and recovered classes progress directly to the susceptible class. However, even with waning some of vaccinated susceptible and recovered individuals might still have some level of protection compared to unvaccinated susceptible individuals. Individuals with pre-existing medical conditions (who can be from both age groups) are not accounted for explicitly. Furthermore, we did not account for specific variants of the virus and the reduced efficacy of vaccines against these variants.

### 4.2. Conclusion

A compartmental model framework for COVID-19 transmission dynamics with different vaccination strategies including vaccination allocation based on two age groups (high or low risk of transmission, severe disease, hospitalization, and death from COVID-19) is developed, parameterized, analyzed rigorously, and used to assess the influence of age structure and vaccine prioritization based on age brackets on the burden of COVID-19 in WA. The results of the study indicate that individuals below 65 years old are the major drivers of the COVID-19 pandemic in WA. This provides one reason for which the reported number of COVID-19 cases in WA is low in comparison to other regions. Therefore, incorporating age-structure in COVID-19 models is important in understanding why the burden of COVID-19 has been low in WA. This result calls for COVID-19 control measures which target individuals below 65 years old as opposed to the elderly as was the case in many high-income countries when the availability of vaccines was limited or during the initial stages of booster vaccine dose administration. Furthermore, the study shows that prioritizing vaccines with higher efficacies will enhance prospects of containing the disease and that the pandemic can be contained in West Africa if 68% of the population is fully vaccinated. However, a drastic change in the current vaccination rate is required for the sub-region to achieve the vaccine-derived herd immunity threshold of 68%. Additionally, the study shows that the emergence of a new more transmissible variant of concern, or easing current control measures in WA could lead to a more devastating wave depending on the easing level. Hence, to curtail the spread of the disease in WA, it is imperative to improve on existing measures (e.g., the vaccination level in conjunction with the WHO recommended NPIs).

## Data Availability

All data produced in the present study are available upon reasonable request to the authors

## Acknowledgements

CNN acknowledges the support of the Simons Foundation (Award #627346) and the National Science Foundation (Grant Number: DMS #2151870). HBT acknowledges the support of the Graduate Research Assistantship in Developing Countries Program (from the International Mathematics Union). In addition, HBT acknowledges support from the Department of Mathematics at the University of Florida for partially funding his visit and for providing him with the resources necessary to carry out this work. Furthermore, HBT acknowledges support from the European Mathematical Society Simons for Africa fellowship Program and Centre d’Excellence Africain en Sciences Mathematiques, Informatique et Applications (CEA-SMIA) Benin, for partially funding his visit to the University of Florida.

## References

[1] E. Dong, H. Du, L. Gardner, An interactive web-based dashboard to track COVID-19 in real time, The Lancet Infectious Diseases (2020).

[2] Africa Centers for Disease Control and Prevention (CDC), Coronavirus disease 2019 (COVID-19), Africa CDC information (Accessed on March 18, 2022). URL https://africacdc.org/{COVID-19}/

[3] A. Boutayeb, The impact of infectious diseases on the development of Africa, Handbook of disease burdens and quality of life measures (2010) 1171.

[4] P. G. Walker, C. Whittaker, O. J. Watson, M. Baguelin, P. Winskill, A. Hamlet, B. A. Djafaara, Z. Cucunubá, D. Olivera Mesa, W. Green, et al., The impact of COVID-19 and strategies for mitigation and suppression in low-and middle-income countries, Science 369 (6502) (2020) 413–422.

[5] T. Roberton, E. D. Carter, V. B. Chou, A. R. Stegmuller, B. D. Jackson, Y. Tam, T. Sawadogo-Lewis, N. Walker, Early estimates of the indirect effects of the COVID-19 pandemic on maternal and child mortality in low-income and middle-income countries: a modelling study, The Lancet Global Health 8 (7) (2020) e901–e908.

[6] Q. Li, X. Guan, P. Wu, X. Wang, L. Zhou, Y. Tong, R. Ren, K. S. Leung, E. H. Lau, J. Y. Wong, et al., Early transmission dynamics in Wuhan, China, of novel coronavirus–infected pneumonia, New England journal of medicine (2020).

[7] V. Ram, L. P. Schaposnik, A modified age-structured sir model for COVID-19 type viruses, Scientific Reports 11 (1) (2021) 1–15.

[8] C. Favas, P. Jarrett, R. Ratnayake, O. J. Watson, F. Checchi, Country differences in transmissibility, age distribution and case-fatality of sars-cov-2: a global ecological analysis, International Journal of Infectious Diseases 114 (2022) 210–218.

[9] H. B. Taboe, K. V. Salako, J. M. Tison, C. N. Ngonghala, R. G. Kakaï, Predicting COVID-19 spread in the face of control measures in West Africa, Mathematical biosciences 328 (2020) 108431.

[10] M. V. Evans, A. Garchitorena, R. J. Rakotonanahary, J. M. Drake, B. Andriamihaja, E. Rajaonarifara, C. N. Ngonghala, B. Roche, M. H. Bonds, J. Rakotonirina, Reconciling model predictions with low reported cases of COVID-19 in Sub-Saharan Africa: insights from madagascar, Global Health Action 13 (1) (2020) 1816044.

[11] N. G. Davies, P. Klepac, Y. Liu, K. Prem, M. Jit, R. M. Eggo, Age-dependent effects in the transmission and control of COVID-19 epidemics, Nature medicine 26 (8) (2020) 1205–1211.

[12] S. R. Simeni Njonnou, N. C. Noumedem Anangmo, F. Kemta Lekpa, D. Noukeu Njinkui, D. Enyama, C. Ngongang Ouankou, E. Vounsia Balti, E. A. Mbono Samba Eloumba, J. R. Moulion Tapouh, S. P. Choukem, The COVID-19 prevalence among children: hypotheses for low infection rate and few severe forms among this age group in Sub-Saharan Africa, Interdisciplinary Perspectives on Infectious Diseases 2021 (2021).

[13] M. Sadarangani, B. A. Raya, J. M. Conway, S. A. Iyaniwura, R. C. Falcao, C. Colijn, D. Coombs, S. Gantt, Importance of COVID-19 vaccine efficacy in older age groups, Vaccine 39 (15) (2021) 2020–2023.

[14] A. T. Huang, B. Garcia-Carreras, M. D. Hitchings, B. Yang, L. C. Katzelnick, S. M. Rattigan, B. A. Borgert, C. A. Moreno, B. D. Solomon, I. Rodriguez-Barraquer, et al., A systematic review of antibody mediated immunity to coronaviruses: antibody kinetics, correlates of protection, and association of antibody responses with severity of disease, MedRxiv (2020).

[15] N. J. Tsagarakis, A. Sideri, P. Makridis, A. Triantafyllou, A. Stamoulakatou, E. Papadogeorgaki, Age-related prevalence of common upper respiratory pathogens, based on the application of the filmarray respiratory panel in a tertiary hospital in greece, Medicine 97 (22) (2018).

[16] K. T. Eames, N. L. Tilston, E. Brooks-Pollock, W. J. Edmunds, Measured dynamic social contact patterns explain the spread of h1n1v influenza, PLoS computational biology 8 (3) (2012) e1002425.

[17] J. Sultana, G. Mazzaglia, N. Luxi, A. Cancellieri, A. Capuano, C. Ferrajolo, C. de Waure, G. Ferlazzo, G. Trifirò, Potential effects of vaccinations on the prevention of COVID-19: rationale, clinical evidence, risks, and public health considerations, Expert review of vaccines 19 (10) (2020) 919–936.

[18] K. M. Bubar, K. Reinholt, S. M. Kissler, M. Lipsitch, S. Cobey, Y. H. Grad, D. B. Larremore, Model-informed COVID-19 vaccine prioritization strategies by age and serostatus, Science 371 (6532) (2021) 916–921.

[19] Africa Centers for Disease Control and Prevention (CDC), COVID-19 vaccination – Africa CDC, Africa CDC information (Accessed: 19-Mar-2022). URL https://africacdc.org/covid-19-vaccination/

[20] World Health Organization (WHO), coronavirus-disease-(covid-19)-vaccines?, WHO information (Accessed: 20-Mar-2022). URL https://www.who.int/news-room/questions-and-answers/item/coronavirus-disease-(covid-19)-vaccines?gclid=CjwKCAjw8sCRBhA6EiwA6_IF4aYLxI_rkHiweUQMDWg3fCDwQWCftXVGvui27WBA63MdksDcU-ujYBoCIisQAvD_BwE&topicsurvey=v8kj13)

[21] P. Adepoju, As COVID-19 vaccines arrive in Africa, Omicron is reducing supply and increasing demand., Nat. med (2021).

[22] L. B. Tlale, L. Gabaitiri, L. K. Totolo, G. Smith, O. Puswane-Katse, E. Ramonna, B. Mothowaeng, J. Tlhakanelo, T. Masupe, G. Rankgoane-Pono, et al., Acceptance rate and risk perception towards the COVID-19 vaccine in Botswana, PloS one 17 (2) (2022) e0263375.

[23] M. Diarra, A. Kebir, C. Talla, A. Barry, J. Faye, D. Louati, L. Opatowski, M. Diop, L. J. White, C. Loucoubar, et al., Non-pharmaceutical interventions and COVID-19 vaccination strategies in Senegal: a modelling study, BMJ global health 7 (2) (2022) e007236.

[24] C. N. Ngonghala, E. Iboi, S. Eikenberry, M. Scotch, C. R. MacIntyre, M. H. Bonds, A. B. Gumel, Mathematical assessment of the impact of non-pharmaceutical interventions on curtailing the 2019 novel coronavirus, Mathematical Biosciences (2020) 108364.

[25] N. M. Ferguson, D. Laydon, G. Nedjati-Gilani, N. Imai, K. Ainslie, M. Baguelin, S. Bhatia, A. Boonyasiri, Z. Cucunubá, G. Cuomo-Dannenburg, et al., Impact of non-pharmaceutical interventions (NPIs) to reduce COVID-19 mortality and healthcare demand, London: Imperial College COVID-19 Response Team, March 16 (2020).

[26] C. N. Ngonghala, E. Iboi, A. B. Gumel, Could masks curtail the post-lockdown resurgence of COVID-19 in the US?, Mathematical Biosciences 329 (2020) 108452.

[27] C. N. Ngonghala, J. R. Knitter, L. Marinacci, M. H. Bonds, A. B. Gumel, Assessing the impact of widespread respirator use in curtailing covid-19 transmission in the usa, Royal Society open science 8 (9) (2021) 210699.

[28] C. N. Ngonghala, P. Goel, D. Kutor, S. Bhattacharyya, Human choice to self-isolate in the face of the COVID-19 pandemic: A game dynamic modelling approach, Journal of Theoretical Biology 521 (2021) 110692. doi:https://doi.org/10.1016/j.jtbi.2021.110692. URL https://www.sciencedirect.com/science/article/pii/S0022519321001144

[29] S. E. Eikenberry, M. Muncuso, E. Iboi, T. Phan, E. Kostelich, Y. Kuang, A. B. Gumel, To mask or not to mask: Modeling the potential for face mask use by the general public to curtail the COVID-19 pandemic, Infectious Disease Modeling 5 (2020) 293–308.

[30] A. B. Gumel, E. A. Iboi, C. N. Ngonghala, G. A. Ngwa, Toward achieving a vaccine-derived herd immunity threshold for covid-19 in the us, Frontiers in Public Health 9 (2021).

[31] Q. Richard, S. Alizon, M. Choisy, M. T. Sofonea, R. Djidjou-Demasse, Age-structured non-pharmaceutical interventions for optimal control of COVID-19 epidemic, PLoS computational biology 17 (3) (2021) e1008776.

[32] B. H. Foy, B. Wahl, K. Mehta, A. Shet, G. I. Menon, C. Britto, Comparing COVID-19 vaccine allocation strategies in India: A mathematical modelling study, International Journal of Infectious Diseases 103 (2021) 431–438.

[33] X. Wang, H. Wu, S. Tang, Assessing age-specific vaccination strategies and post-vaccination reopening policies for COVID-19 control using seir modeling approach, medRxiv (2021).

[34] W. O. Kermack, A. G. McKendrick, A contribution to the mathematical theory of epidemics, Proceedings of the Royal Society of London. Series A, Containing papers of a mathematical and physical character 115 (772) (1927) 700–721.

[35] R. Pastorino, A. M. Pezzullo, L. Villani, F. A. Causio, C. Axfors, D. G. Contopoulos-Ioannidis, S. Boccia, J. P. Ioannidis, Change in age distribution of COVID-19 deaths with the introduction of COVID-19 vaccination, Environmental research 204 (2022) 112342.

[36] O. Diekmann, J. A. P. Heesterbeek, J. A. Metz, On the definition and the computation of the basic reproduction ratio *R*_0_ in models for infectious diseases in heterogeneous populations, Journal of Mathematical Biology 28 (4) (1990) 365–382.

[37] P. van den Driessche, J. Watmough, Reproduction numbers and sub-threshold endemic equilibria for compartmental models of disease transmission, Mathematical Biosciences 180 (1-2) (2002) 29–48.

[38] T. Sahel, W. A. C. S. (SWAC), Tackling the coronavirus (COVID-19): West african perspectives, COVID-19 in West Africa, SWAC information (Accessed on May 07, 2022). URL https://www.oecd.org/swac/coronavirus-west-africa/

[39] A. A. King, M. Domenech de Cellès, F. M. Magpantay, P. Rohani, Avoidable errors in the modelling of outbreaks of emerging pathogens, with special reference to Ebola, Proceedings of the Royal Society B: Biological Sciences 282 (1806) (2015) 20150347.

[40] A. C. Davison, D. V. Hinkley, Bootstrap methods and their application, no. 1, Cambridge university press, 1997.

[41] S. Marino, I. B. Hogue, C. J. Ray, D. E. Kirschner, A methodology for performing global uncertainty and sensitivity analysis in systems biology, Journal of theoretical biology 254 (1) (2008) 178–196.

[42] Centers for Disease Control and Prevention, Different COVID-19 vaccines, CDC information (Accessed on January 25, 2021). URL https://www.cdc.gov/coronavirus/2019-ncov/vaccines/different-vaccines.html

[43] M. J. Keeling, T. D. Hollingsworth, J. M. Read, The efficacy of contact tracing for the containment of the 2019 novel coronavirus (COVID-19)., medRxiv (2020).

[44] M. Mbow, B. Lell, S. P. Jochems, B. Cisse, S. Mboup, B. G. Dewals, A. Jaye, A. Dieye, M. Yazdanbakhsh, COVID-19 in Africa: Dampening the storm?, Science 369 (6504) (2020) 624–626.

[45] The World Bank, Population ages 65 and above (% of total population) - Sub-Saharan Africa, WorldBank (Assessed on june 29, 2022). URL https://data.worldbank.org/indicator/SP.POP.65UP.TO.ZS?locations=ZG

[46] K. B. Blyuss, Y. N. Kyrychko, Effects of latency and age structure on the dynamics and containment of COVID-19, Journal of Theoretical Biology 513 (2021) 110587.

[47] S. M. Moghadas, M. C. Fitzpatrick, P. Sah, A. Pandey, A. Shoukat, B. H. Singer, A. P. Galvani, The implications of silent transmission for the control of COVID-19 outbreaks, Proceedings of the National Academy of Sciences (2020). arXiv:https://www.pnas.org/content/early/2020/07/02/2008373117.full. pdf, doi:10.1073/pnas.2008373117. URL https://www.pnas.org/content/early/2020/07/02/2008373117

[48] L. C. Tindale, J. E. Stockdale, M. Coombe, E. S. Garlock, W. Y. V. Lau, M. Saraswat, L. Zhang, D. Chen, J. Wallinga, C. Colijn, Evidence for transmission of COVID-19 prior to symptom onset, eLife 9 (2020) e57149.

[49] O. S. Ilesanmi, A. Akande, A. A. Afolabi, Overcoming COVID-19 in West African countries: is herd immunity an option?, The Pan African Medical Journal 35 (Suppl 2) (2020).

[50] D. M. Altmann, D. C. Douek, R. J. Boyton, What policy makers need to know about COVID-19 protective immunity, The Lancet 395 (10236) (2020) 1527–1529.

[51] M. O. Afolabi, O. Wariri, Y. Saidu, A. Otu, S. A. Omoleke, B. Ebenso, A. Adebiyi, M. Ooko, B. O. Ahinkorah, E. K. Ameyaw, et al., Tracking the uptake and trajectory of COVID-19 vaccination coverage in 15 west african countries: an interim analysis, BMJ global health 6 (12) (2021) e007518.

[52] H. Ritchie, E. Ortiz-Ospina, D. Beltekian, E. Mathieu, J. Hasell, B. Macdonald, C. Giattino, M. Roser, Coronavirus (COVID-19) vaccinations, Statistics and Research, Our World in Data (Accessed on January 24, 2021).

[53] Centers for Disease Control and Prevention (CDC), Cdc expands eligibility for covid-19 booster shots to all adults, CDC information (Accessed on February 22, 2022). URL https://www.cdc.gov/media/releases/2021/s1119-booster-shots.html

[54] M.-M. Mehrabi Nejad, F. Moosaie, H. Dehghanbanadaki, A. Haji Ghadery, M. Shabani, M. Tabary, A. Aryannejad, S. SeyedAlinaghi, N. Rezaei, Immunogenicity of COVID-19 mRNA vaccines in immunocompromised patients: a systematic review and meta-analysis, European journal of medical research 27 (1) (2022) 1–13.

