## Supplementary material for "Impact of age-structure and vaccine prioritization on COVID-19 in West Africa": Online supplementary information

### Online supplementary information (SI)

#### 1 Brief description of model variables and parameters

Table S1: Brief descriptions of the variables of the basic model (Model 1)

| Variable | Description |
|---|---|
| $S_u$ | Unvaccinated susceptible individuals, i.e., unvaccinated individuals who have not contracted the virus. |
| $S_v$ | Fully vaccinated susceptible individuals, i.e., vaccinated individuals who have not contracted the virus. |
| $E$ | Latent individuals, i.e., individuals who have acquired the virus but have not started spreading it. |
| $I_p$ | Presymptomatic infectious individuals. These are infected individuals who start transmitting the virus without exhibiting any disease symptom and before the end of the incubation period. |
| $I_a$ | Asymptomatic infectious individuals, i.e., infectious individuals who do not exhibit clinical disease symptoms at the end of the incubation period. |
| $I_i$ | Symptomatic infectious individuals, i.e., infectious individuals who exhibit clinical disease symptoms at the end of the incubation period. |
| $I_c$ | Confirmed (reported) cases, i.e., individuals who test positive to COVID-19. |
| $I_h$ | Individuals who are hospitalized because of COVID-19. |
| $R$ | Recovered individuals with temporary immunity against the virus. |

Table S2: Brief descriptions of the parameters of the basic model (S.1).

| Parameter | Definition or description |
|---|---|
| $\Lambda$ | Recruitment rate (all recruitment are into the unvaccinated susceptible class). |
| $\mu$ | Natural death rate in each of the compartments. |
| $\omega_r$ | Natural immunity waning rate ($1/\omega_r$ is the average duration of natural immunity). |
| $\omega_v$ | Vaccine-derived immunity waning rate ($1/\omega_v$ is the average duration of vaccine immunity). |
| $\xi$ | Vaccination rate of individuals from the unvaccinated susceptible and recovered classes. |
| $\varepsilon$ | The efficacy of vaccines in preventing vaccinated individuals from becoming infected. |
| $\tau_a$ | Positivity rate for latent, presymptomatic, and asymptomatic infectious individuals. |
| $\tau_i$ | Positivity rate for symptomatic infectious individuals. |
| $r$ | Proportion of presymptomatic infectious individuals who become symptomatic. |
| $\phi_c(\phi_i)$ | Hospitalization rate of individuals from the symptomatic infectious (confirmed) class. |
| $\delta_i(\delta_c)(\delta_h)$ | Disease-induced death rate for individuals from the $I_i$ ($I_c$) ($I_h$) class. |
| $(1/\sigma_e)(1/\sigma_p)$ | Average length of time spent in the latent (presymptomatic infectious) class. |
| $\gamma_a(\gamma_c)(\gamma_i)(\gamma_h)$ | Recovery rate of individuals from the $I_a$ $(I_c)$ $(I_i)$ $(I_h)$ class. |
| $\beta_a(\beta_c)(\beta_p)(\beta_i)(\beta_h)$ | Effective transmission rate for individuals from the $I_a$ ($I_c$) ($I_p$) ($I_i$) ($I_h$) class. |

Table S3: Brief descriptions of the variables of the vaccine-structured model (Model 2).

| Variable | Description |
|---|---|
| $S_u$ | Unvaccinated susceptible individuals, i.e., unvaccinated individuals who have not contracted the virus. |
| $E_u$ | Latent unvaccinated individuals, i.e., individuals who have acquired the virus but have not started spreading it and are not vaccinated. |
| $I_{pu}$ | Presymptomatic infectious unvaccinated individuals. These are infected unvaccinated individuals who start transmitting the virus without exhibiting any disease symptom and before the end of the incubation period. |
| $I_{au}$ | Asymptomatic infectious unvaccinated individuals, i.e., infectious individuals who do not exhibit clinical disease symptoms at the end of the incubation period and are style not vaccinated . |
| $I_{iu}$ | Symptomatic infectious unvaccinated individuals, i.e., infectious individuals who exhibit clinical disease symptoms at the end of the incubation period. |
| $I_{cu}$ | Unvaccinated confirmed (reported) cases, i.e., individuals who test positive to COVID-19 and was not vaccinated. |
| $I_{hu}$ | Unvaccinated individuals who are hospitalized because of COVID-19. |
| $R_u$ | Recovered unvaccinated individuals with temporary immunity against the virus but who were not vaccinated. |
| $S_v$ | Fully vaccinated susceptible individuals, i.e., vaccinated individuals who have not contracted the virus. |
| $E_v$ | Latent vaccinated individuals, i.e., individuals who have acquired the virus but have not started spreading it and are vaccinated. |
| $I_{pv}$ | Presymptomatic infectious vaccinated individuals. These are infected vaccinated individuals who start transmitting the virus without exhibiting any disease symptom and before the end of the incubation period. |
| $I_{av}$ | Asymptomatic infectious vaccinated individuals, i.e., infectious individuals who do not exhibit clinical disease symptoms at the end of the incubation period and are style vaccinated . |
| $I_{iv}$ | Symptomatic infectious vaccinated individuals, i.e., infectious individuals who exhibit clinical disease symptoms at the end of the incubation period. |
| $I_{cv}$ | Vaccinated confirmed (reported) cases, i.e., individuals who test positive to COVID-19 and was vaccinated. |
| $I_{hv}$ | Vaccinated individuals who are hospitalized because of COVID-19. |
| $R_v$ | Recovered vaccinated individuals with temporary immunity against the virus. |

Table S4: Brief descriptions of the parameters of the vaccine-structured model (Model 2).

| Parameter | Definition or description |
|---|---|
| $\Lambda$ | Recruitment rate (all recruitment are into the unvaccinated susceptible class). |
| $\mu$ | Natural death rate in each of the compartments. |
| $\omega_{ru}$ | Natural immunity waning rate ($1/\omega_{ru}$ is the average duration of natural immunity). |
| $\omega_{sv}$ | Vaccine-derived immunity waning rate ($1/\omega_{sv}$ is the average duration of vaccine immunity). |
| $\xi$ | Vaccination rate of individuals . |
| $\varepsilon$ | The efficacy of vaccines in preventing vaccinated individuals from becoming infected. |
| $\tau_a$ | Positivity rate for latent, presymptomatic, and asymptomatic infectious individuals. |
| $\tau_i$ | Positivity rate for symptomatic infectious individuals. |
| $r$ | Proportion of presymptomatic infectious individuals who become symptomatic. |
| $\pi$ | Reduction in transmission due to vaccination of the infectious individual. |
| $\pi_h$ | Reduction in hospitalization due to vaccination of the infected individual. |
| $\phi_{cm}(\phi_{im})$ | Hospitalization rate of individuals m (m=u for unvaccinated and m=v for vaccinated) from the $m$ symptomatic infectious (confirmed) class . |
| $\delta_{im}(\delta_{cm})(\delta_{hm})$ | Disease-induced death rate for $m$ individuals from the $I_{im}$ ($I_{cm}$) ($I_{hm}$) class. |
| $(1/\sigma_e)(1/\sigma_p)$ | Average length of time spent in the latent (presymptomatic infectious) class. |
| $\gamma_{am}(\gamma_{cm})(\gamma_{im})(\gamma_{hm})$ | Recovery rate of $m$ individuals from the $I_{am}$ ($I_{cm}$) ($I_{im}$) ($I_{hm}$) class. |
| $\beta_{amn}(\beta_{cmn})(\beta_{pmn})(\beta_{imn})(\beta_{hmn})$ | Effective transmission rate from a $jm, j \in \{p, a, i, c, h\}$ infectious class to an $n$ susceptible class) with $m, n \in \{u, v\}$ . |

Table S5: Brief descriptions of the variables of the vaccine-age-structured model (Model 3).

| Variable | Description |
|---|---|
| $S_{uk}$ | Unvaccinated susceptible individuals in age-Group k (k=1 for youths and k= 2 for adults). |
| $E_{uk}$ | Latent unvaccinated individuals in age-Group k . |
| $I_{puk}$ | Presymptomatic infectious unvaccinated individuals in age-Group k . |
| $I_{auk}$ | Asymptomatic infectious unvaccinated individuals in age-Group k . |
| $I_{iuk}$ | Symptomatic infectious unvaccinated individuals in age-Group k . |
| $I_{cuk}$ | Unvaccinated confirmed (reported) cases in age-Group k |
| $I_{huk}$ | Unvaccinated individuals in age-Group k who are hospitalized because of COVID-19. |
| $R_{uk}$ | Recovered unvaccinated individuals in age-Group k with temporary immunity against the virus . |
| $S_{vk}$ | Fully vaccinated susceptible individuals in age-Group k. |
| $E_{vk}$ | Latent vaccinated individuals in age-Group k. |
| $I_{pvk}$ | Presymptomatic infectious vaccinated individuals in age-Group k. |
| $I_{avk}$ | Asymptomatic infectious vaccinated individuals in age-Group k. |
| $I_{ivk}$ | Symptomatic infectious vaccinated individualsin age-Group k. |
| $I_{cvk}$ | Vaccinated confirmed (reported) cases in age-Group k. |
| $I_{hvk}$ | Vaccinated individuals in age-Group k who are hospitalized because of COVID-19. |
| $R_{vk}$ | Recovered vaccinated individuals in age-Group k with temporary immunity against the virus. |

Table S6: Brief descriptions of the parameters of the vaccine-age-structured model (Model 3).

| **Parameter** | **Definition or description** |
|---|---|
| $\Lambda$ | Recruitment rate . |
| $\mu$ | Natural death rate in each of the compartments. |
| $\rho$ | Maturation rate of the youth (e.g., the rate at which youths become adults) |
| $\omega_{ruk}$ | Natural immunity waning rate ($1/\omega_{ruk}$ is the average duration of natural immunity of individual in age-group k). |
| $\omega_{svk}$ | Vaccine-derived immunity waning rate ($1/\omega_{svk}$ is the average duration of vaccine immunity) in age-group k. |
| $\xi_k$ | Vaccination rate of individuals in age-group k . |
| $\varepsilon$ | The efficacy of vaccines in preventing vaccinated individuals from becoming infected. |
| $\tau_a$ | Positivity rate for latent, presymptomatic, and asymptomatic infectious individuals. |
| $\tau_{ik}$ | Positivity rate for symptomatic infectious individuals in age-group k. |
| $r_k$ | Proportion of presymptomatic infectious individuals in age-group k who become symptomatic. |
| $\pi$ | Reduction in transmission due to vaccination of the infectious individual. |
| $\pi_h$ | Reduction in hospitalization due to vaccination of the infected individual. |
| $\phi_{cmk}(\phi_{imk})$ | Hospitalization rate of individuals m in age-group k from the $m$ symptomatic infectious (confirmed) class . |
| $\delta_{imk}(\delta_{cmk})(\delta_{hmk})$ | Disease-induced death rate for $mk$ individuals from the $I_{imk}$ ($I_{cmk}$) ($I_{hmk}$) class. |
| $(1/\sigma_e)(1/\sigma_p)$ | Average length of time spent in the latent (presymptomatic infectious). |
| $\gamma_{amk}(\gamma_{cmk})(\gamma_{imk})(\gamma_{hmk})$ | Recovery rate of $mk$ individuals from the $I_{amk}$ ($I_{cmk}$) ($I_{imk}$) ($I_{hmk}$). |
| $\beta_{amnk}(\beta_{cmnk})(\beta_{pmnk})$ $(\beta_{imnk})(\beta_{hmnk})$ | Effective transmission rate from a $jmk, j \in \{p, a, i, c, h\}$ infectious class to an $nk$ susceptible class) with $m, n \in \{u, v\}$ . |

#### 2 Model 1 without vaccination (Model 0)

The equations of the basic model without vaccination (mentioned in section 3.2 of the main paper) are as follow:

$$\begin{aligned}
\dot{S}_u &= \Lambda + \omega_r\, R - \lambda\, S_u - \mu\, S_u, \\
\dot{E} &= \lambda\, S_u - (\tau_a + \sigma_e + \mu)\, E, \\
\dot{I}_p &= \sigma_e\, E - (\tau + \sigma_p + \mu)\, I_p, \\
\dot{I}_i &= r\, \sigma_p\, I_p - (\tau_i + \phi_i + \gamma_i + \delta_i + \mu)\, I_i, \\
\dot{I}_a &= (1 - r)\, \sigma_p\, I_p - (\tau_a + \gamma_a + \mu)\, I_a, \\
\dot{I}_c &= \tau_a\, (E + I_p + I_a) + \tau_i I_i - (\gamma_c + \phi_c + \delta_c + \mu)\, I_c, \\
\dot{I}_h &= \phi_i\, I_i + \phi_c I_c - (\gamma_h + \delta_h + \mu)\, I_h, \\
\dot{R} &= \gamma_i\, I_i + \gamma_a\, I_a + \gamma_h\, I_h + \gamma_c I_c - (\mu + \omega_r)\, R.
\end{aligned} \tag{S.1}$$

#### 3 Matrices used to calculate the reproduction number for each model

In this section, we provide the matrices used to compute the reproduction. It is worth to recall that the reproduction number is the spectral radius of the Next generation matrix given by the product of new infection matrix $F$ and the inverse of the transition matrix $V$ [1]. In other words, the control reproduction number $\mathcal{R}_c$, is given by :

$$\mathcal{R}_c = \varrho(F \times V^{-1}) \tag{S.1}$$

In (S.1), $\varrho$ is a function that return the spectral radius of the next generation matrix $F \times V^-1$. The basic reproduction number is computed the same way but in absence of control measures (the vaccination related or any other control measure's parameters will be replaced by zero in each matrix).

##### 3.1 Model 0

Let $F_0$ and $V_0$ be respectively the matrix of new infection and transition matrix for the model without vaccination. $F_0$ and $V_0$ are given by:

$$F_0 = \begin{bmatrix} 0 & \beta_p & \beta_i & \beta_a & \beta_c & \beta_h \\ 0 & 0 & 0 & 0 & 0 & 0 \\ 0 & 0 & 0 & 0 & 0 & 0 \\ 0 & 0 & 0 & 0 & 0 & 0 \\ 0 & 0 & 0 & 0 & 0 & 0 \\ 0 & 0 & 0 & 0 & 0 & 0 \end{bmatrix}, \; V_0 = \begin{bmatrix} A_e & 0 & 0 & 0 & 0 & 0 \\ -\sigma_e & A_p & 0 & 0 & 0 & 0 \\ 0 & -r\sigma_p & A_i & 0 & 0 & 0 \\ 0 & -(1-r)\,\sigma_p & 0 & A_a & 0 & 0 \\ -\tau_a & -\tau_a & -\tau_i & -\tau_a & A_c & 0 \\ 0 & 0 & -\phi_i & 0 & -\phi_c & A_h \end{bmatrix}$$

With
$A_e = \tau_a + \sigma_e + \mu,\; A_p = \tau_a + \sigma_p + \mu,\; A_i = \tau_i + \phi_i + \gamma_i + \delta_i + \mu,\; A_a = \tau_a + \gamma_a + \mu,\; A_c = \gamma_c + \phi_c + \delta_c + \mu,$
$A_h = \gamma_h + \delta_h + \mu.$
The control reproduction number of model 0 is obtained by replacing matrices $F$ and $V$ in (S.1) by respectively matrices $F_0$ and $V_0$.

##### 3.2 Model 1

Let $F_1$ and $V_1$ be respectively the matrix of new infection and transition matrix for the model without vaccination. $F_1$ and $V_1$ are given by:

$$F_1 = \frac{S_u^* + (1-\epsilon)S_v^*}{S_u^* + S_v^*} \begin{bmatrix} 0 & \beta_p & \beta_i & \beta_a & \beta_c & \beta_h \\ 0 & 0 & 0 & 0 & 0 & 0 \\ 0 & 0 & 0 & 0 & 0 & 0 \\ 0 & 0 & 0 & 0 & 0 & 0 \\ 0 & 0 & 0 & 0 & 0 & 0 \\ 0 & 0 & 0 & 0 & 0 & 0 \end{bmatrix}, \; V_1 = \begin{bmatrix} A_e & 0 & 0 & 0 & 0 & 0 \\ -\sigma_e & A_p & 0 & 0 & 0 & 0 \\ 0 & -r\sigma_p & A_i & 0 & 0 & 0 \\ 0 & -(1-r)\,\sigma_p & 0 & A_a & 0 & 0 \\ -\tau_a & -\tau_a & -\tau_i & -\tau_a & A_c & 0 \\ 0 & 0 & -\phi_i & 0 & -\phi_c & A_h \end{bmatrix}$$

With

$$S_u^* = \frac{\Lambda(\mu + \omega_v)}{\mu(\mu + \omega_v + \xi)},\; S_v^* = \frac{\Lambda\xi}{\mu(\mu + \omega_v + \xi)}$$

And
$A_e = \tau_a + \sigma_e + \mu,\; A_p = \tau_a + \sigma_p + \mu,\; A_i = \tau_i + \phi_i + \gamma_i + \delta_i + \mu,\; A_a = \tau_a + \gamma_a + \mu,\; A_c = \gamma_c + \phi_c + \delta_c + \mu,$
$A_h = \gamma_h + \delta_h + \mu.$
The reproduction number of model 1 is obtained by replacing matrices $F$ and $V$ in (S.1) by respectively matrices $F_1$ and $V_1$. The basic reproduction number is obtained by replacing by zero in the expression of $R_c$ all the control measures related parameters. The herd immunity is calculated using the Eq.3.3 in the main paper.

#### 3.3 Model2

Let $F_2$ and $V_2$ be respectively the matrix of new infection and transition matrix for the model without vaccination. $F_2$ and $V_2$ are given by:

$$F_2 = \begin{bmatrix} F_{211} & F_{212} & F_{213} \\ F_{221} & F_{222} & F_{223} \\ F_{231} & F_{232} & F_{233} \end{bmatrix}, \; V_2 = \begin{bmatrix} V_{211} & V_{212} & V_{213} \\ V_{221} & V_{222} & V_{223} \\ V_{231} & V_{232} & V_{233} \end{bmatrix}$$

Where $F_{211} = \frac{S_u^*}{N^*}\begin{bmatrix} 0 & \beta_{puu} & \beta_{iuu} & \beta_{auu} \\ 0 & 0 & 0 & 0 \\ 0 & 0 & 0 & 0 \\ 0 & 0 & 0 & 0 \end{bmatrix}$, $F_{212} = \frac{S_u^*}{N^*}\begin{bmatrix} \beta_{cuu} & \beta_{huu} & 0 & \beta_{pvu} \\ 0 & 0 & 0 & 0 \\ 0 & 0 & 0 & 0 \\ 0 & 0 & 0 & 0 \end{bmatrix}$

$$F_{213} = \frac{S_u^*}{N^*}\begin{bmatrix} \beta_{ivu} & \beta_{avu} & \beta_{cvu} & \beta_{hvu} \\ 0 & 0 & 0 & 0 \\ 0 & 0 & 0 & 0 \\ 0 & 0 & 0 & 0 \end{bmatrix}, \; F_{221} = \frac{S_v^*}{N^*}\begin{bmatrix} 0 & 0 & 0 & 0 \\ 0 & 0 & 0 & 0 \\ 0 & \beta_{puv} & \beta_{iuv} & \beta_{auv} \\ 0 & 0 & 0 & 0 \end{bmatrix},$$

$$F_{222} = \frac{S_v^*}{N^*}\begin{bmatrix} 0 & 0 & 0 & 0 \\ 0 & 0 & 0 & 0 \\ \beta_{cuv} & \beta_{huv} & 0 & \beta_{pvv} \\ 0 & 0 & 0 & 0 \end{bmatrix}, \; F_{223} = \frac{S_v^*}{N^*}\begin{bmatrix} 0 & 0 & 0 & 0 \\ 0 & 0 & 0 & 0 \\ \beta_{ivv} & \beta_{avv} & \beta_{cvv} & \beta_{hvv} \\ 0 & 0 & 0 & 0 \end{bmatrix}$$

$$F_{231} = F_{232} = F_{233} = \begin{bmatrix} 0 & 0 & 0 & 0 \\ 0 & 0 & 0 & 0 \\ 0 & 0 & 0 & 0 \\ 0 & 0 & 0 & 0 \end{bmatrix} \; V_{211} = \begin{bmatrix} A_u & 0 & 0 & 0 \\ -\sigma_e & A_{pu} & 0 & 0 \\ 0 & -r\sigma_p & A_{iu} & 0 \\ 0 & -(1-r)\,\sigma_p & 0 & A_{au} \end{bmatrix},$$

$$V_{212} = V_{213} = V_{223} = \begin{bmatrix} 0 & 0 & 0 & 0 \\ 0 & 0 & 0 & 0 \\ 0 & 0 & 0 & 0 \\ 0 & 0 & 0 & 0 \end{bmatrix}, \; V_{221} = \begin{bmatrix} -\tau_a & \tau_a & -\tau_i & -\tau_a \\ 0 & 0 & -\phi_{iu} & 0 \\ -\xi & 0 & 0 & 0 \\ 0 & -\xi & 0 & 0 \end{bmatrix}, V_{222} = \begin{bmatrix} A_{cu} & 0 & 0 & 0 \\ -\phi_{cu} & A_{hu} & 0 & 0 \\ 0 & 0 & A_v & 0 \\ 0 & 0 & -\sigma_e & A_{pv} \end{bmatrix}$$

$$V_{231} = \begin{bmatrix} 0 & 0 & 0 & 0 \\ 0 & 0 & 0 & -\xi \\ 0 & 0 & 0 & 0 \\ 0 & 0 & 0 & 0 \end{bmatrix}, \; V_{232} = \begin{bmatrix} 0 & 0 & 0 & -r\sigma_p \\ 0 & 0 & 0 & -(1-r)\,\sigma_p \\ 0 & 0 & -\tau_a & -\tau_a \\ 0 & 0 & 0 & 0 \end{bmatrix}, V_{233} = \begin{bmatrix} A_{iv} & 0 & 0 & 0 \\ 0 & A_{av} & 0 & 0 \\ -\tau_i & -\tau_a & A_{cv} & 0 \\ -\phi_{iv} & 0 & -\phi_{cv} & A_{hv} \end{bmatrix}$$

With
$A_u = \xi + \tau_a + \sigma_e + \mu,\; A_{pu} = \xi + \tau_a + \sigma_p + \mu,\; A_{iu} = \tau_i + \phi_{iu} + \gamma_{iu} + \delta_{iu} + \mu,\; A_{au} = \xi + \tau_a + \gamma_{au} + \mu,$
$A_{cu} = \gamma_c + \phi_{cu} + \delta_{cu} + \mu,\; A_{hu} = \gamma_{hu} + \delta_{hu} + \mu,\; A_v = \tau_a + \sigma_e + \mu,\; A_{pv} = \tau_a + \sigma_p + \mu,\; A_{iv} = \tau_i + \phi_{iv} + \gamma_{iv} + \delta_{iv} + \mu,$
$A_{av} = \tau_a + \gamma_{av} + \mu,\; A_{cv} = \gamma_{cv} + \phi_{cv} + \delta_{cv} + \mu,\; A_{hv} = \gamma_{hv} + \delta_{hv} + \mu.$

The disease-free equilibrium of the model is $(S_u^*, E_u^*, I_{pu}^*, I_{iu}^*, I_{au}^*, I_{cu}^*, I_{hu}^*, R_u^*, S_v^*, E_v^*, I_{pv}^*, I_{iv}^*, I_{av}^*, I_{cv}^*, I_{hv}^*, R_v^*)$ = $\left(\frac{(\mu+\omega_{sv})\Lambda}{\mu(\mu+\xi+\omega_{sv})}, 0, 0, 0, 0, 0, 0, 0, \frac{\Lambda\xi}{\mu(\mu+\xi+\omega_{sv})}, 0, 0, 0, 0, 0, 0, 0\right)$. The reproduction number of model 2 is obtained by replacing matrices $F$ and $V$ in (S.1) by respectively matrices $F_2$ and $V_2$. The basic reproduction number is obtained by replacing by zero in the expression of $R_c$ all the control measures related parameters. The herd immunity is calculated using the Eq.3.3 in the main paper.

#### 3.4 Model3

Let $F_3$ and $V_3$ be respectively the matrix of new infection and transition matrix for the model without vaccination. $F_3$ and $V_3$ are given by:

$$F_3 = \begin{bmatrix} F_{311} & F_{312} & F_{313} \\ F_{321} & F_{322} & F_{323} \\ F_{331} & F_{332} & F_{333} \end{bmatrix}, V_3 = \begin{bmatrix} V_{311} & V_{312} & V_{313} \\ V_{321} & V_{322} & V_{323} \\ V_{331} & V_{332} & V_{333} \end{bmatrix}$$

Where

$$F_{311} = \begin{bmatrix} 0 & S^*_{u1}\beta_{pu1} & S^*_{u1}\beta_{iu1} & S^*_{u1}\beta_{au1} & S^*_{u1}\beta_{cu1} & S^*_{u1}\beta_{hu1} & 0 & S^*_{u1}\beta_{pu1}(1-\pi) \\ 0 & 0 & 0 & 0 & 0 & 0 & 0 & 0 \\ 0 & 0 & 0 & 0 & 0 & 0 & 0 & 0 \\ 0 & 0 & 0 & 0 & 0 & 0 & 0 & 0 \\ 0 & 0 & 0 & 0 & 0 & 0 & 0 & 0 \\ 0 & 0 & 0 & 0 & 0 & 0 & 0 & 0 \\ 0 & X_v S^*_{v1}\beta_{pu1} & X_v S^*_{v1}\beta_{iu1} & X_v S^*_{v1}\beta_{au1} & X_v S^*_{v1}\beta_{cu1} & X_v S^*_{v1}\beta_{hu1} & 0 & X_v S^*_{v1}\beta_{pu1}(1-\pi) \\ 0 & 0 & 0 & 0 & 0 & 0 & 0 & 0 \end{bmatrix}$$

$$F_{312} = \begin{bmatrix} S_{u1}\beta_{iu1}R_t & S_{u1}\beta_{au1}R_t & S_{u1}\beta_{cu1}R_t & S_{u1}\beta_{hu1}R_t & 0 & \frac{S_{u1}\beta_{pu12}}{2} & \frac{S_{u1}\beta_{iu12}}{2} & \frac{S_{u1}\beta_{au12}}{2} \\ 0 & 0 & 0 & 0 & 0 & 0 & 0 & 0 \\ 0 & 0 & 0 & 0 & 0 & 0 & 0 & 0 \\ 0 & 0 & 0 & 0 & 0 & 0 & 0 & 0 \\ 0 & 0 & 0 & 0 & 0 & 0 & 0 & 0 \\ 0 & 0 & 0 & 0 & 0 & 0 & 0 & 0 \\ S_{v1}\beta_{iu1}X_x & S_{v1}\beta_{au1}X_x & S_{v1}\beta_{cu1}X_x & S_{v1}\beta_{hu1}X_x & 0 & \frac{S_{v1}\beta_{pu12}X_v}{2} & \frac{S_{v1}\beta_{iu12}X_v}{2} & \frac{S_{v1}\beta_{au12}X_v}{2} \\ 0 & 0 & 0 & 0 & 0 & 0 & 0 & 0 \end{bmatrix}$$

$$F_{313} = \begin{bmatrix} \frac{S_{u1}\beta_{cu12}}{2} & \frac{S_{u1}\beta_{hu12}}{2} & 0 & S_{u1}\beta_{pu12}R_{t2} & S_{u1}\beta_{iu12}R_{t2} & S_{u1}\beta_{au12}R_{t2} & S_{u1}\beta_{cu12}R_{t2} & S_{u1}\beta_{hu12}R_{t2} \\ 0 & 0 & 0 & 0 & 0 & 0 & 0 & 0 \\ 0 & 0 & 0 & 0 & 0 & 0 & 0 & 0 \\ 0 & 0 & 0 & 0 & 0 & 0 & 0 & 0 \\ 0 & 0 & 0 & 0 & 0 & 0 & 0 & 0 \\ 0 & 0 & 0 & 0 & 0 & 0 & 0 & 0 \\ \frac{S_{v1}\beta_{cu12}X_v}{2} & \frac{S_{v1}\beta_{hu12}X_v}{2} & 0 & S_{v1}\beta_{pu12}X_r & S_{v1}\beta_{iu12}X_r & S_{v1}\beta_{au12}X_r & S_{v1}\beta_{cu12}X_r & S_{v1}\beta_{hu12}X_r \\ 0 & 0 & 0 & 0 & 0 & 0 & 0 & 0 \end{bmatrix}$$

$$F_{321} = \begin{bmatrix} 0 & 0 & 0 & 0 & 0 & 0 & 0 & 0 \\ 0 & 0 & 0 & 0 & 0 & 0 & 0 & 0 \\ 0 & 0 & 0 & 0 & 0 & 0 & 0 & 0 \\ 0 & 0 & 0 & 0 & 0 & 0 & 0 & 0 \\ 0 & S_{u2}\beta_{pu12} & S_{u2}\beta_{iu12} & S_{u2}\beta_{au12} & S_{u2}\beta_{cu12} & S_{u2}\beta_{hu12} & 0 & S_{u2}\beta_{pu12}R_t \\ 0 & 0 & 0 & 0 & 0 & 0 & 0 & 0 \\ 0 & 0 & 0 & 0 & 0 & 0 & 0 & 0 \\ 0 & 0 & 0 & 0 & 0 & 0 & 0 & 0 \end{bmatrix}$$

$$F_{322} = \begin{bmatrix} 0 & 0 & 0 & 0 & 0 & 0 & 0 & 0 \\ 0 & 0 & 0 & 0 & 0 & 0 & 0 & 0 \\ 0 & 0 & 0 & 0 & 0 & 0 & 0 & 0 \\ 0 & 0 & 0 & 0 & 0 & 0 & 0 & 0 \\ S_{u2}\beta_{iu12}R_t & S_{u2}\beta_{au12}R_t & S_{u2}\beta_{cu12}R_t & S_{u2}\beta_{hu12}R_t & 0 & S_{u2}\beta_{pu2} & S_{u2}\beta_{iu2} & S_{u2}\beta_{au2} \\ 0 & 0 & 0 & 0 & 0 & 0 & 0 & 0 \\ 0 & 0 & 0 & 0 & 0 & 0 & 0 & 0 \\ 0 & 0 & 0 & 0 & 0 & 0 & 0 & 0 \end{bmatrix}$$

$$F_{323}=\begin{bmatrix} 0&0&0&0&0&0&0&0\\ 0&0&0&0&0&0&0&0\\ 0&0&0&0&0&0&0&0\\ 0&0&0&0&0&0&0&0\\ S_{u2}\beta_{cu2}&S_{u2}\beta_{hu2}&0&S_{u2}\beta_{pu2}R_t&S_{u2}\beta_{iu2}R_t&S_{u2}\beta_{au2}R_t&S_{u2}\beta_{cu2}R_t&S_{u2}\beta_{hu2}R_t\\ 0&0&0&0&0&0&0&0\\ 0&0&0&0&0&0&0&0\\ 0&0&0&0&0&0&0&0 \end{bmatrix}$$

$$F_{331}=\begin{bmatrix} 0&0&0&0&0&0&0&0\\ 0&0&0&0&0&0&0&0\\ 0&S_{v2}\beta_{pu12}X_v&S_{v2}\beta_{iu12}X_v&S_{v2}\beta_{au12}X_v&S_{v2}\beta_{cu12}X_v&S_{v2}\beta_{hu12}X_v&0&S_{v2}\beta_{pu12}X_x\\ 0&0&0&0&0&0&0&0\\ 0&0&0&0&0&0&0&0\\ 0&0&0&0&0&0&0&0\\ 0&0&0&0&0&0&0&0\\ 0&0&0&0&0&0&0&0 \end{bmatrix}$$

$$F_{332}=\begin{bmatrix} 0&0&0&0&0&0&0&0\\ 0&0&0&0&0&0&0&0\\ S_{v2}\beta_{iu12}X_x&S_{v2}\beta_{au12}X_x&S_{v2}\beta_{cu12}X_x&S_{v2}\beta_{hu12}X_x&0&S_{v2}\beta_{pu12}X_v&S_{v2}\beta_{iu2}X_v&S_{v2}\beta_{au2}X_v\\ 0&0&0&0&0&0&0&0\\ 0&0&0&0&0&0&0&0\\ 0&0&0&0&0&0&0&0\\ 0&0&0&0&0&0&0&0\\ 0&0&0&0&0&0&0&0 \end{bmatrix}$$

$$F_{333}=\begin{bmatrix} 0&0&0&0&0&0&0&0\\ 0&0&0&0&0&0&0&0\\ S_{v2}\beta_{cu2}X_v&S_{v2}\beta_{hu2}X_v&0&S_{v2}\beta_{pu2}X_x&S_{v2}\beta_{iu2}X_x&S_{v2}\beta_{au2}X_x&S_{v2}\beta_{cu2}X_x&S_{v2}\beta_{hu2}X_x\\ 0&0&0&0&0&0&0&0\\ 0&0&0&0&0&0&0&0\\ 0&0&0&0&0&0&0&0\\ 0&0&0&0&0&0&0&0\\ 0&0&0&0&0&0&0&0 \end{bmatrix}$$

With $X_v=(1-\varepsilon)$, $X_r=X_vR_{t2}$,$X_x=X_vR_t$, $R_t=1-\pi$ and $R_{t2}=\frac{r_t}{2}-\frac{1}{2}$
And

$$V_{311}=\begin{bmatrix} A_{eu1}&0&0&0&0&0&0&0\\ -\sigma_e&A_{pu1}&0&0&0&0&0&0\\ 0&-r_1\sigma_p&A_{iu1}&0&0&0&0&0\\ 0&\sigma_p\left(r_1-1\right)&0&A_{au1}&0&0&0&0\\ -\tau_a&-\tau_a&-\tau_{i1}&-\tau_a&A_{cu1}&0&0&0\\ 0&0&-\phi_{iu1}&0&-\phi_{cu1}&A_{hu1}&0&0\\ -\xi_1&0&0&0&0&0&A_{ev1}&0\\ 0&-\xi_1&0&0&0&0&-\sigma_e&A_{pv1} \end{bmatrix}$$

$$V_{312} = V_{313} = V_{323} = \begin{bmatrix} 0 & 0 & 0 & 0 & 0 & 0 & 0 & 0 \\ 0 & 0 & 0 & 0 & 0 & 0 & 0 & 0 \\ 0 & 0 & 0 & 0 & 0 & 0 & 0 & 0 \\ 0 & 0 & 0 & 0 & 0 & 0 & 0 & 0 \\ 0 & 0 & 0 & 0 & 0 & 0 & 0 & 0 \\ 0 & 0 & 0 & 0 & 0 & 0 & 0 & 0 \\ 0 & 0 & 0 & 0 & 0 & 0 & 0 & 0 \\ 0 & 0 & 0 & 0 & 0 & 0 & 0 & 0 \end{bmatrix}$$

$$V_{321} = \begin{bmatrix} 0 & 0 & 0 & 0 & 0 & 0 & 0 & -r_1\sigma_p \\ 0 & 0 & 0 & -\xi_1 & 0 & 0 & 0 & \sigma_p\left(r_1 - 1\right) \\ 0 & 0 & 0 & 0 & 0 & 0 & -\tau_a & -\tau_a \\ 0 & 0 & 0 & 0 & 0 & 0 & 0 & 0 \\ -\rho & 0 & 0 & 0 & 0 & 0 & 0 & 0 \\ 0 & -\rho & 0 & 0 & 0 & 0 & 0 & 0 \\ 0 & 0 & -\rho & 0 & 0 & 0 & 0 & 0 \\ 0 & 0 & 0 & -\rho & 0 & 0 & 0 & 0 \end{bmatrix}$$

$$V_{322} = \begin{bmatrix} A_{iv1} & 0 & 0 & 0 & 0 & 0 & 0 & 0 \\ 0 & A_{av1} & 0 & 0 & 0 & 0 & 0 & 0 \\ -\tau_{i1} & \tau_a & A_{cv1} & 0 & 0 & 0 & 0 & 0 \\ -\phi_{iv1} & 0 & -\phi_{cv1} & A_{hv1} & 0 & 0 & 0 & 0 \\ 0 & 0 & 0 & 0 & A_{eu2} & 0 & 0 & 0 \\ 0 & 0 & 0 & 0 & \sigma_e & A_{pu2} & 0 & 0 \\ 0 & 0 & 0 & 0 & 0 & -r_2\sigma_p & A_{iu2} & 0 \\ 0 & 0 & 0 & 0 & 0 & \sigma_p\left(r_2 - 1\right) & 0 & A_{au2} \end{bmatrix}$$

$$V_{331} = \begin{bmatrix} 0 & 0 & 0 & 0 & -\rho & 0 & 0 & 0 \\ 0 & 0 & 0 & 0 & 0 & -\rho & 0 & 0 \\ 0 & 0 & 0 & 0 & 0 & 0 & -\rho & 0 \\ 0 & 0 & 0 & 0 & 0 & 0 & 0 & -\rho \\ 0 & 0 & 0 & 0 & 0 & 0 & 0 & 0 \\ 0 & 0 & 0 & 0 & 0 & 0 & 0 & 0 \\ 0 & 0 & 0 & 0 & 0 & 0 & 0 & 0 \\ 0 & 0 & 0 & 0 & 0 & 0 & 0 & 0 \end{bmatrix}, \; V_{332} = \begin{bmatrix} 0 & 0 & 0 & 0 & -\tau_a & -\tau_a & -\tau_{i2} & -\tau_a \\ 0 & 0 & 0 & 0 & 0 & 0 & -\phi_{iu2} & 0 \\ 0 & 0 & 0 & 0 & -\xi_2 & 0 & 0 & 0 \\ 0 & 0 & 0 & 0 & 0 & -\xi_2 & 0 & 0 \\ -\rho & 0 & 0 & 0 & 0 & 0 & 0 & 0 \\ 0 & -\rho & 0 & 0 & 0 & 0 & 0 & -\xi_2 \\ 0 & 0 & -\rho & 0 & 0 & 0 & 0 & 0 \\ 0 & 0 & 0 & -\rho & 0 & 0 & 0 & 0 \end{bmatrix},$$

$$V_{333} = \begin{bmatrix} A_{cu2} & 0 & 0 & 0 & 0 & 0 & 0 & 0 \\ -\phi_{cu2} & A_{hu2} & 0 & 0 & 0 & 0 & 0 & 0 \\ 0 & 0 & A_{ev2} & 0 & 0 & 0 & 0 & 0 \\ 0 & 0 & -\sigma_e & A_{pv2} & 0 & 0 & 0 & 0 \\ 0 & 0 & 0 & -r_2\sigma_p & A_{iv2} & 0 & 0 & 0 \\ 0 & 0 & 0 & \sigma_p\left(r_2 - 1\right) & 0 & A_{av2} & 0 & 0 \\ 0 & 0 & -\tau_a & -\tau_a & -\tau_{i2} & -\tau_a & A_{cv2} & 0 \\ 0 & 0 & 0 & 0 & -\phi_{iv2} & 0 & -\phi_{cv2} & A_{hv2} \end{bmatrix}$$

With
$A_{eu1} = \mu_1 + \sigma_e + \tau_a + \xi_1 + \rho$ , $A_{pu1} = \mu_1 + \sigma_p + \tau_a + \xi_1 + \rho$, $A_{iu1} = \delta_{iu1} + \gamma_{iu1} + \mu_1 + \phi_{iu1} + \rho + \tau_{i1}$,
$A_{au1} = \gamma_{au1} + \mu_1 + \rho + \tau_a + \xi_1$ , $A_{cu1} = \delta_{cu1} + \gamma_{cu1} + \mu_1 + \phi_{cu1} + \rho$, $A_{hu1} = \delta_{hu1} + \gamma_{hu1} + \mu_1 + \rho$,
$A_{ev1} = \mu_1 + \rho + \sigma_e + \tau_a$ , $A_{pv1} = \mu_1 + \rho + \sigma_p + \tau_a$, $A_{iv1} = \delta_{iv1} + \gamma_{iv1} + \mu_1 + \phi_{iv1} + \rho + \tau_{i1}$,
$A_{av1} = \gamma_{av1} + \mu_1 + \rho + \tau_a$ , $A_{cv1} = \delta_{cv1} + \gamma_{cv1} + \mu_1 + \phi_{cv1} + \rho$, $A_{hv1} = \delta_{hv1} + \gamma_{hv1} + \mu_1 + \rho$,
$A_{eu2} = \mu_2 + \sigma_e + \tau_a + \xi_2$, $A_{pu2} = \mu_2 + \sigma_p + \tau_a + \xi_2$, $A_{iu2} = \delta_{iu2} + \gamma_{iu2} + \mu_2 + \phi_{iu2} + \tau_{i2}$,
$A_{au2} = \gamma_{au2} + \mu_2 + \tau_a + \xi_2$, $A_{cu2} = \delta_{cu2} + \gamma_{cu2} + \mu_2 + \phi_{cu2}$, $A_{hu2} = \delta_{hu2} + \gamma_{hu2} + \mu_2$,

$A_{ev2} = \mu_2 + \sigma_e + \tau_a,\ A_{pv2} = \mu_2 + \sigma_p + \tau_a,\ A_{iv2} = \delta_{iv2} + \gamma_{iv2} + \mu_2 + \phi_{iv2} + \tau_{i2},\ A_{av2} = \gamma_{av2} + \mu_2 + \tau_a,$ $A_{cv2} = \delta_{cv2} + \gamma_{cv2} + \mu_2 + \phi_{cv2},\ A_{hv2} = \delta_{hv2} + \gamma_{hv2} + \mu_2.$

The disease-free equilibrium of the model is $(S^*_{u1}, E^*_{u1}, I^*_{pu1}, I^*_{iu1}, I^*_{au1}, I^*_{cu1}, I^*_{hu1}, R^*_{u1}, S^*_{v1}, E^*_{v1}, I^*_{pv1}, I^*_{iv1}, I^*_{av1}, I^*_{cv1}, I^*_{hv1}, R^*_{v1}, S^*_{u2}, E^*_{u2}, I^*_{pu2}, I^*_{iu2}, I^*_{au2}, I^*_{cu2}, I^*_{hu2}, R^*_{u2}, S^*_{v2}, E^*_{v2}, I^*_{pv2}, I^*_{iv2}, I^*_{av2}, I^*_{cv2}, I^*_{hv2}, R^*_{v2})$ = $(S^*_{u1}, 0, 0, 0, 0, 0, 0, 0, S^*_{v1}, 0, 0, 0, 0, 0, 0, 0, S^*_{u2}, 0, 0, 0, 0, 0, 0, 0, S^*_{v2}, 0, 0, 0, 0, 0, 0, 0).$

Where
$S^*_{u1} = \frac{(\mu_1+\omega_{sv1}+\rho)\Lambda}{\mu_1^2+(2\rho+\xi_1+\omega_{sv1})\mu_1+(\rho-w_{sv1}+\omega_{sv1})\xi_1+\rho(\omega_{sv1}+\rho)},\ S^*_{v2} = \frac{\Lambda((\xi_2+\mu_2)\xi_1+\xi_2(\mu_1+\omega_{sv1}+\rho))\rho}{((\rho+\mu_1-w_{sv1}+\omega_{sv1})\xi_1+(\rho+\mu_1)(\mu_1+\omega_{sv1}+\rho))\mu_2(\mu_2+\xi_2+\omega_{sv2})},$
$S^*_{v1} = \frac{\Lambda\xi_1}{(\rho+\mu_1-w_{sv1}+\omega_{sv1})\xi_1+(\rho+\mu_1)(\mu_1+\omega_{sv1}+\rho)},\ S^*_{u2} = \frac{((\mu_2+\omega_{sv2})\rho+(\mu_2+\omega_{sv2})\mu_1+(\xi_1+\omega_{sv1})\omega_{sv2}+\mu_2\omega_{sv1})\Lambda\rho}{\left(\rho^2+(2\mu_1+\xi_1+\omega_{sv1})\rho+\mu_1^2+(\xi_1+\omega_{sv1})\mu_1-\xi_1(w_{sv1}-\omega_{sv1})\right)\mu_2(\mu_2+\xi_2+\omega_{sv2})}$
The reproduction number of model 3 is obtained by replacing matrices $F$ and $V$ in (S.1) by respectively matrices $F_3$ and $V_3$. The basic reproduction number is obtained by replacing by zero in the expression of $R_c$ all the control measures related parameters. The herd immunity is calculated using the Eq.3.3 in the main paper.

### 4 Fixed and estimated baseline parameter values

#### 4.1 Fixed parameters

Table S7: Fixed baseline parameter values of the models. Apart from the vaccine efficacy ($\epsilon$), and the proportions $\theta$, which are dimensionless, all the other parameters have units of *per* day.

| Parameter | Value | Source | Parameter | Value | Source | Parameter | Value | Source |
|---|---|---|---|---|---|---|---|---|
| $\Lambda$ | $1.94 \times 10^4$ | [2] * | $\rho$ | $0.024/365$ | [6]* | $r_1$ | 0.1 | [10]* |
| $\mu$ | $1/21900$ | [2] * | $\sigma_e$ | $1/3$ | [7] | $r_2$ | 0.3 | [10] |
| $\phi_i$ | $1/6$ | [3] | $\sigma_p$ | $1/2$ | [7] | $\xi_1$ | $2.3 \times 10^{-6}$ | [9]* |
| $\phi_c$ | $1/3$ | [3] | $\epsilon$ | 0.674 | [8, 9] | $\xi_2$ | $9.4 \times 10^{-6}$ | [9]* |
| $\omega_r$ | $12/3285$ | [3–5] | $\xi$ | $1.17 \times 10^{-5}$ | [9]* | $\pi$ | 0.76 | [11] |
| $\omega_{sv}$ | $12/3285$ | [3–5] | $r$ | 0.20 | [10] | $\pi_h$ | 0.71 | [12] |

*: *Adapted based on the epidemiological or demographic data on the research area.*

#### 4.2 Estimated parameters

Table S8: Estimated (fitted) baseline parameter values and confidence intervals (CIs) for the model 0 using COVID-19 confirmed case data for the WA for Wave 1 (...). The unit of each of the estimated community transmission rate is *per* day.

(a) Wave 1 (from 02/28/2020 to 10/25/2020).

| Parameters | Value | 95% CI |
|---|---|---|
| $\beta_p$ | 0.1464127 | [0.1395931, 0.2207476] |
| $\beta_a$ | 0.2580704 | [0.1846186, 0.2894376] |
| $\beta_i$ | 0.1923896 | [0.1564342, 0.2597074] |
| $\beta_h$ | 0.0737172 | [0.0603023, 0.1000000] |
| $\delta_i$ | 0.0001826 | [0.0001266, 0.0001863] |
| $\delta_h$ | 0.0000934 | [0.0000932, 0.0002743] |
| $\gamma_i$ | 0.2212030 | [0.1859248, 0.2500000] |
| $\gamma_a$ | 0.3071068 | [0.3093787, 0.3333322] |
| $\gamma_h$ | 0.0667357 | [0.0666667, 0.1257132] |
| $\tau$ | 0.0000101 | [0.0000100, 0.0000786] |
| $\tau_i$ | 0.0010234 | [0.0001005, 0.0013643] |
| $\gamma_c$ | 0.2641549 | [0.2476517, 0.2916661] |
| $\beta_c$ | 0.0737172 | [0.0603023, 0.10000000] |

(b) Wave 2 (from 10/26/2020 to 05/31/2021)

| Parameters | Value | 95% CI |
|---|---|---|
| $\beta_p$ | 0.1203940 | [0.0701311, 0.1311305] |
| $\beta_a$ | 0.3215022 | [0.3085735, 0.4556457] |
| $\beta_i$ | 0.2824220 | [0.2625455, 0.4462928] |
| $\beta_h$ | 0.0479575 | [0.0027682, 0.0999991] |
| $\delta_i$ | 0.0001388 | [0.0001755, 0.0001863] |
| $\delta_h$ | 0.0001416 | [0.0000932, 0.0002775] |
| $\gamma_i$ | 0.2494546 | [0.2061641, 0.2499999] |
| $\gamma_a$ | 0.3333333 | [0.3170441, 0.3333239] |
| $\gamma_h$ | 0.0666667 | [0.0666667, 0.2778075] |
| $\tau$ | 0.0001047 | [0.0000100, 0.0001160] |
| $\tau_i$ | 0.0002050 | [0.0001001, 0.0016928] |
| $\gamma_c$ | 0.2913940 | [0.2616041, 0.2916619] |
| $\beta_c$ | 0.0479575 | [0.0027682, 0.0999991] |

Table S9: Estimated (fitted) baseline parameter values and confidence intervals (CIs) for the model 1 using COVID-19 confirmed case data for the WA for Wave 3 (from 5/31/2021 to 11/14/2021) and Wave 4 (from 11/14/2021 to 3/14/2022). The unit of each of the estimated community transmission rate is *per* day.

(a) Wave 3 (from 06/01/2021 to 11/14/2021).

| Parameters | Value | 95% CI |
|---|---|---|
| $\beta_p$ | 0.0930451 | [0.0700002, 0.2048215] |
| $\beta_a$ | 0.3526031 | [0.2219806, 0.4567197] |
| $\beta_i$ | 0.0476395 | [0.0001467, 0.0916129] |
| $\beta_h$ | 0.0440261 | [0.0000141, 0.0700000] |
| $\delta_i$ | 0.0001584 | [0.0000621, 0.0001863] |
| $\delta_h$ | 0.0002792 | [0.0000932, 0.0002795] |
| $\gamma_i$ | 0.2311843 | [0.0500000, 0.2499998] |
| $\gamma_a$ | 0.2325097 | [0.1783571, 0.3256428] |
| $\gamma_h$ | 0.0689244 | [0.0666667, 0.3326895] |
| $\tau$ | 0.0000859 | [0.0000100, 0.0001092] |
| $\tau_i$ | 0.0001000 | [0.0001000, 0.0017553] |
| $\gamma_c$ | 0.2318470 | [0.1141786, 0.2878213] |
| $\beta_c$ | 0.0440261 | [0.0000141, 0.0700000] |
| $\delta_c$ | 0.0001396 | [0.0000466, 0.0001398] |

(b) Wave 4 (11/15/2021 to 3/14/2022).

| Parameters | Value | 95% CI |
|---|---|---|
| $\beta_p$ | 0.2460421 | [0.0726911, 0.4646603] |
| $\beta_a$ | 0.6131425 | [0.2623308, 0.7531487] |
| $\beta_i$ | 0.1055269 | [0.0000019, 0.2099985] |
| $\beta_h$ | 0.0005141 | [0.0000000, 0.0010000] |
| $\delta_i$ | 0.0001843 | [0.0000621, 0.0001863] |
| $\delta_h$ | 0.0002795 | [0.0000932, 0.0002800] |
| $\gamma_i$ | 0.2499997 | [0.0500077, 0.2500000] |
| $\gamma_a$ | 0.3205945 | [0.1375376, 0.3333333] |
| $\gamma_h$ | 0.1140679 | [0.0666673, 0.3333333] |
| $\tau$ | 0.0000187 | [0.0000100, 0.0000616] |
| $\tau_i$ | 0.0007247 | [0.0001001, 0.0008338] |
| $\gamma_c$ | 0.2852971 | [0.0937726, 0.2916667] |
| $\beta_c$ | 0.0005141 | [0.0000000, 0.0010000] |
| $\delta_c$ | 0.0001398 | [0.0000466, 0.0001400] |

Table S10: Estimated (fitted) baseline parameter values and confidence intervals (CIs) for the model 2 using COVID-19 confirmed case data for the WA for Wave 3 (from 06/01/2021 to 11/14/2021) and Wave 4 (from 11/15/2021 to 3/14/2022). The unit of each of the estimated community transmission rate is *per* day.

(a) Wave 3 (from 06/01/2021 to 11/14/2021).

| Parameters | Value | 95% CI |
|---|---|---|
| $\beta_{pu}$ | 0.1321999 | [0.0041315, 0.2224804] |
| $\beta_{iu}$ | 0.0253787 | [0.0000002, 0.2396418] |
| $\beta_{au}$ | 0.2774134 | [0.1469887, 0.4213462] |
| $\beta_{cu}$ | 0.0002833 | [0.0000005, 0.0006277] |
| $\beta_{hu}$ | 0.0000425 | [0.0000000, 0.0001000] |
| $\delta_{iu}$ | 0.0002184 | [0.0000932, 0.0002795] |
| $\delta_{hu}$ | 0.0001897 | [0.0000311, 0.0003730] |
| $\gamma_{iu}$ | 0.2474801 | [0.1250141, 0.3333333] |
| $\gamma_{au}$ | 0.2910105 | [0.1885021, 0.3333325] |
| $\gamma_{hu}$ | 0.1331148 | [0.0588235, 0.1666666] |
| $\tau_a$ | 0.0000623 | [0.0000001, 0.0001390] |
| $\tau_i$ | 0.0010097 | [0.0000100, 0.0023516] |
| $\omega_{ru}$ | 0.0012032 | [0.0009132, 0.0013699] |
| $\omega_{sv}$ | 0.0009548 | [0.0007828, 0.0010959] |
| $\omega_{rv}$ | 0.0008301 | [0.0006849, 0.0009132] |
| $\delta_{iv}$ | 0.0000671 | [0.0000286, 0.0000858] |
| $\delta_{hv}$ | 0.0000582 | [0.0000095, 0.0001145] |
| $\delta_{cu}$ | 0.0000949 | [0.0000155, 0.0001865] |
| $\delta_{cv}$ | 0.0000291 | [0.0000048, 0.0000573] |
| $\gamma_{iv}$ | 0.2474801 | [0.1250141, 0.3333333] |
| $\gamma_{av}$ | 0.2910105 | [0.1885021, 0.3333325] |
| $\gamma_{hv}$ | 0.1331148 | [0.0588235, 0.1666666] |
| $\gamma_{cu}$ | 0.2692453 | [0.1567581, 0.3333329] |
| $\gamma_{cv}$ | 0.2692453 | [0.1567581, 0.3333329] |
| $R_v$ | 1.1535090 | [0.63363334, 1.7672816] |

(b) Wave 4 (11/15/2021 to 3/14/2022).

| Parameters | Value | 95% CI |
|---|---|---|
| $\beta_{pu}$ | 0.2345009 | [0.1053475, 0.4094053] |
| $\beta_{iu}$ | 0.0382829 | [0.0000001, 0.290939] |
| $\beta_{au}$ | 0.4322110 | [0.1652186, 0.5974481] |
| $\beta_{cu}$ | 0.0004577 | [0.0000265, 0.0007452] |
| $\beta_{hu}$ | 0.0000498 | [0.0000000, 0.0001000] |
| $\delta_{iu}$ | 0.0002332 | [0.0000933, 0.0002795] |
| $\delta_{hu}$ | 0.0002224 | [0.0000311, 0.0003729] |
| $\gamma_{iu}$ | 0.2631594 | [0.1453736, 0.3333101] |
| $\gamma_{au}$ | 0.3028938 | [0.1793899, 0.3333321] |
| $\gamma_{hu}$ | 0.1023382 | [0.0588235, 0.166664] |
| $\tau_a$ | 0.0000112 | [0.0000001, 0.0000596] |
| $\tau_i$ | 0.0008899 | [0.0000484, 0.0012267] |
| $\omega_{ru}$ | 0.0010461 | [0.0009132, 0.0013699] |
| $\omega_{sv}$ | 0.0008875 | [0.0007828, 0.0010959] |
| $\omega_{rv}$ | 0.0007847 | [0.0006849, 0.0009132] |
| $\delta_{iv}$ | 0.0000716 | [0.0000287, 0.0000858] |
| $\delta_{hv}$ | 0.0000683 | [0.0000095, 0.0001145] |
| $\delta_{cu}$ | 0.0001112 | [0.0000155, 0.0001864] |
| $\delta_{cv}$ | 0.0000341 | [0.0000048, 0.0000572] |
| $\gamma_{iv}$ | 0.2631594 | [0.1453736, 0.3333101] |
| $\gamma_{av}$ | 0.3028938 | [0.1793899, 0.3333321] |
| $\gamma_{hv}$ | 0.1023382 | [0.0588235, 0.166664] |
| $\gamma_{cu}$ | 0.2830266 | [0.1623817, 0.3333211] |
| $\gamma_{cv}$ | 0.2830266 | [0.1623817, 0.3333211] |
| $R_v$ | 1.8562172 | [1.0486517, 2.7690552] |

Table S11: Estimated (fitted) baseline parameter values and confidence intervals (CIs) for the model 3 using COVID-19 confirmed case data for the WA for Wave 3 (from 06/01/2021 to 11/14/2021) and Wave 4 (from 11/15/2021 to 03/14/2022). The unit of each of the estimated community transmission rate is *per* day.

(a) Wave 3 (06/01/2021 to 11/14/2021).

| Parameters | Value | 95% CI |
|---|---|---|
| $\beta_{pu1}$ | 0.1290311 | [0.0750735, 0.2491480] |
| $\beta_{iu1}$ | 0.0002160 | [0.0001000, 0.0703188] |
| $\beta_{au1}$ | 0.4123497 | [0.3118220, 0.4407702] |
| $\beta_{cu1}$ | 0.0003473 | [0.0000109, 0.0005876] |
| $\beta_{hu1}$ | 0.0000830 | [0.0000217, 0.0001000] |
| $\beta_{pu2}$ | 0.2938881 | [0.0578108, 0.4999541] |
| $\beta_{iu2}$ | 0.2914213 | [0.0415157, 0.4999224] |
| $\beta_{au2}$ | 0.0371803 | [0.0163465, 0.0499969] |
| $\beta_{cu2}$ | 0.0003608 | [0.0000490, 0.0004999] |
| $\beta_{hu2}$ | 0.0000007 | [0.0000000, 0.0000010] |
| $\beta_{au12}$ | 0.0000498 | [0.0000000, 0.0001000] |
| $\beta_{pu12}$ | 0.1083342 | [0.0158560, 0.3087768] |
| $\beta_{iu12}$ | 0.0000022 | [0.0000000, 0.0000050] |
| $\beta_{hu12}$ | 0.0866503 | [0.0080868, 0.0999900] |
| $\beta_{cu12}$ | 0.2397093 | [0.0152751, 0.4999887] |
| $\gamma_{iu1}$ | 0.3049307 | [0.0557600, 0.8828517] |
| $\gamma_{iv1}$ | 0.3049307 | [0.0557600, 0.8828517] |
| $\gamma_{au1}$ | 0.1906077 | [0.1226403, 0.2819773] |
| $\gamma_{av1}$ | 0.1906077 | [0.1226403, 0.2819773] |
| $\gamma_{hu1}$ | 0.1365819 | [0.1000017, 0.2673167] |
| $\gamma_{hv1}$ | 0.1365819 | [0.1000017, 0.2673167] |
| $\gamma_{cu1}$ | 0.2477692 | [0.0892002, 0.5824145] |
| $\gamma_{cv1}$ | 0.2477692 | [0.0892002, 0.5824145] |
| $\gamma_{iu2}$ | 0.0765028 | [0.0002704, 0.2007806] |
| $\gamma_{iv2}$ | 0.0765028 | [0.0002704, 0.2007806] |
| $\gamma_{au2}$ | 0.0437072 | [0.0298282, 0.4742942] |
| $\gamma_{av2}$ | 0.0437072 | [0.0298282, 0.4742942] |
| $\gamma_{hu2}$ | 0.0724150 | [0.0700007, 0.3196793] |
| $\gamma_{hv2}$ | 0.0724150 | [0.0700007, 0.3196793] |
| $\gamma_{cu2}$ | 0.0601050 | [0.0150493, 0.3375374] |
| $\gamma_{cv2}$ | 0.0601050 | [0.0150493, 0.3375374] |
| $\tau_{i2}$ | 0.0021902 | [0.0012838, 0.0032051] |
| $\tau_{i1}$ | 0.0019188 | [0.0000003, 0.0052153] |
| $\tau_{a}$ | 0.0000010 | [0.0000010, 0.0000433] |

(b) Wave 4 (11/15/2021 to 3/14/2022).

| Parameters | Value | 95% CI |
|---|---|---|
| $\beta_{pu1}$ | 0.2431314 | [0.0298496, 0.3510959] |
| $\beta_{iu1}$ | 0.1871759 | [0.0058773, 0.6080411] |
| $\beta_{au1}$ | 0.5913658 | [0.4407630, 0.8809824] |
| $\beta_{cu1}$ | 0.0050113 | [0.0027583, 0.0089634] |
| $\beta_{hu1}$ | 0.0006430 | [0.0000000, 0.0010000] |
| $\beta_{pu2}$ | 0.0052986 | [0.0001025, 0.0099998] |
| $\beta_{iu2}$ | 0.0278770 | [0.0000001, 0.0998676] |
| $\beta_{au2}$ | 0.0251694 | [0.0000330, 0.0499970] |
| $\beta_{cu2}$ | 0.0002362 | [0.0000001, 0.0004999] |
| $\beta_{hu2}$ | 0.0000005 | [0.0000000, 0.0000010] |
| $\beta_{au12}$ | 0.0000352 | [0.0000000, 0.0000999] |
| $\beta_{pu12}$ | 0.0640447 | [0.0000001, 0.3012572] |
| $\beta_{iu12}$ | 0.0000019 | [0.0000000, 0.0000050] |
| $\beta_{hu12}$ | 0.0273801 | [0.0000004, 0.0979258] |
| $\beta_{cu12}$ | 0.0681128 | [0.0000000, 0.4554436] |
| $\gamma_{iu1}$ | 0.2843679 | [0.0481989, 0.7308239] |
| $\gamma_{iv1}$ | 0.2843679 | [0.0481989, 0.7308239] |
| $\gamma_{au1}$ | 0.2342220 | [0.0815646, 0.3543681] |
| $\gamma_{av1}$ | 0.2342220 | [0.0815646, 0.3543681] |
| $\gamma_{hu1}$ | 0.0878150 | [0.0100013, 0.2892208] |
| $\gamma_{hv1}$ | 0.0878150 | [0.0100013, 0.2892208] |
| $\gamma_{cu1}$ | 0.2592949 | [0.0648817, 0.5425960] |
| $\gamma_{cv1}$ | 0.2592949 | [0.0648817, 0.5425960] |
| $\gamma_{iu2}$ | 0.2327523 | [0.0000003, 0.8741621] |
| $\gamma_{iv2}$ | 0.2327523 | [0.0000003, 0.8741621] |
| $\gamma_{au2}$ | 0.3496527 | [0.0013103, 0.9368965] |
| $\gamma_{av2}$ | 0.3496527 | [0.0013103, 0.9368965] |
| $\gamma_{hu2}$ | 0.1774915 | [0.0700002, 0.3199986] |
| $\gamma_{hv2}$ | 0.0022687 | [0.0012493, 0.0033314] |
| $\gamma_{cu2}$ | 0.2912025 | [0.0006553, 0.9055293] |
| $\gamma_{cv2}$ | 0.2912025 | [0.0006553, 0.9055293] |
| $\tau_{i2}$ | 0.0000104 | [0.0000000, 0.0000503] |
| $\tau_{i1}$ | 0.0023365 | [0.0000795, 0.0056351] |
| $\tau_{a}$ | 0.0000104 | [0.0000000, 0.0000503] |

### 5 Investigating the impact of vaccine efficacy and vaccine coverage on the burden of COVID-19 in WA

Table S12: coverage thresholds for corresponding vaccine efficacies (Extracted from Fig. 6 of the main paper)

| **Efficacy** | $f_v$ **model 1** | $f_v$ **model 2** | $f_v$ **model 3** |
|---|---|---|---|
| 0.76 | 0.7475 | 0.6364 | 0.6061 |
| 0.669 | 0.8485 | 0.7273 | 0.6869 |
| 0.79 | 0.7172 | 0.6061 | 0.5758 |
| 0.51 | 1.0000 | 0.9495 | 0.8990 |
| 0.64 | 0.8889 | 0.7576 | 0.7172 |
| 0.941 | 0.6970 | 0.5152 | 0.4848 |
| 0.6738 | 0.8384 | 0.7273 | 0.6768 |

### 6 Investigating the impact of full vaccination on the dynamics of COVID-19 in WA.

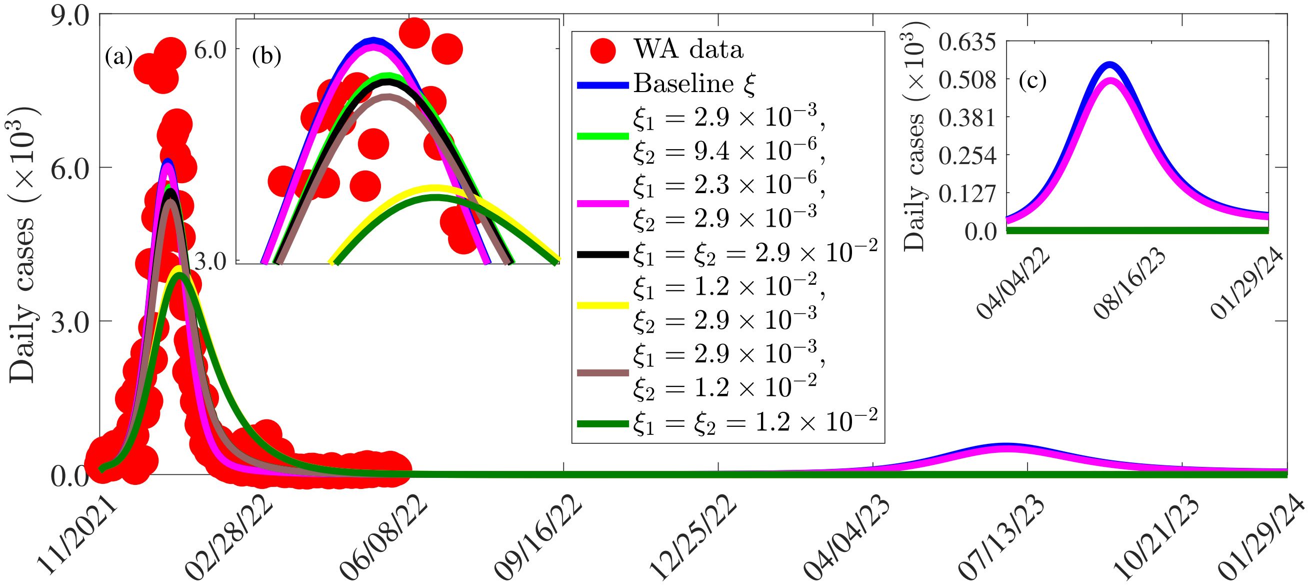


Figure S1: Impact of fully Vaccination rate per age of the dynamic of COVID19 daily case in WA.

### References


[1] O. Diekmann, J. A. P. Heesterbeek, and J. A. Metz, "On the definition and the computation of the basic reproduction ratio $R_0$ in models for infectious diseases in heterogeneous populations," Journal of Mathematical Biology **28**, 365–382 (1990).

[2] C. N. Ngonghala, E. Iboi, and A. B. Gumel, "Could masks curtail the post-lockdown resurgence of COVID-19 in the US?" Mathematical Biosciences **329**, 108452 (2020).

[3] J. Seow, C. Graham, B. Merrick, S. Acors, S. Pickering, K. J. Steel, O. Hemmings, A. O'Byrne, N. Kouphou, R. P. Galao, et al., "Longitudinal observation and decline of neutralizing antibody responses in the three months following SARS-CoV-2 infection in humans," Nature microbiology **5**, 1598–1607 (2020).

[4] B. Curley, "How long does immunity from COVID-19 vaccination last?" Healthline (Accessed on July 25, 2021) (2021).

[5] J. M. Dan, J. Mateus, Y. Kato, K. M. Hastie, E. D. Yu, C. E. Faliti, A. Grifoni, S. I. Ramirez, S. Haupt, A. Frazier, et al., "Immunological memory to sars-cov-2 assessed for up to 8 months after infection," Science **371**, eabf4063 (2021).

[6] Mariam Saleh, "Growth rate of the population of africa in 2020, by country," statista (Accessed on March 18, 2022).
Online Version

[7] N. M. Linton, T. Kobayashi, Y. Yang, K. Hayashi, A. R. Akhmetzhanov, S.-m. Jung, B. Yuan, R. Kinoshita, and H. Nishiura, "Incubation period and other epidemiological characteristics of 2019 novel coronavirus infections with right truncation: a statistical analysis of publicly available case data," Journal of Clinical Medicine **9**, 538 (2020).

[8] World Health Organization, "COVID-19 advice for the public: Getting vaccinated," WHO (Assessed on june 29, 2022).
Online Version

[9] Africa Centers for Disease Control and Prevention (CDC), "COVID-19 vaccination – Africa cdc," Africa CDC information (Accessed: 19-Mar-2022).
Online Version

[10] R. Subramanian, Q. He, and M. Pascual, "Quantifying asymptomatic infection and transmission of COVID-19 in New York City using observed cases, serology, and testing capacity," Proceedings of the National Academy of Sciences **118**, e2019716118 (2021).

[11] D. W. Eyre, D. Taylor, M. Purver, D. Chapman, T. Fowler, K. B. Pouwels, A. S. Walker, and T. E. Peto, "Effect of covid-19 vaccination on transmission of alpha and delta variants," New England Journal of Medicine (2022).

[12] Centers for Disease Control and Prevention (CDC), "COVID-19 vaccinations in the United States," CDC COVID-19 data tracker (Accessed on February 16, 2022).
Online Version
